# Chart review and genetic validation of electronic medical record dementia diagnoses in VA: The impact of CMS data

**DOI:** 10.64898/2026.07.14.26358063

**Authors:** Mark W. Logue, Sophia O. Lee, Margaret Gillis, Rui Zhang, Monica T. Ly, David Marra, Francesca V. Lopez, Julie Lynch, Matthew S. Panizzon, Debby W. Tsuang, Richard L. Hauger, The MVP Cognitive Decline and Dementia During Aging Working Group, VA Million Veteran Program, Victoria C. Merritt

## Abstract

**Background:** International Classification of Diseases (ICD) codes are often used in epidemiological studies to track disease rates over time.

**Objective:** This evaluation of ICD-code-based algorithms for electronic medical record (EMR) studies of Alzheimer’s disease (AD) and related dementias (ADRD) examines the impact of incorporating Centers for Medicare and Medicaid (CMS) data as an additional source of diagnostic and treatment information in Department of Veterans Affairs (VA) EMR studies.

**Methods:** We performed a chart review of 100 VA Million Veteran Program (MVP) participants to evaluate algorithm performance. We also assessed genetic associations across algorithms in a large MVP cohort (n=396k).

**Results:** Adding CMS data increased the number of detected cases, sensitivity, and positive predictive value, but decreased specificity and negative predictive value. Genetic analyses showed that broader (ADRD/dementia) algorithms with just VA data performed similarly to narrow (AD-focused) algorithms incorporating both VA and CMS ICD codes. Additionally, narrow AD algorithms based solely on VA data yielded the highest ORs, indicating the largest proportion of late-onset AD cases.

**Conclusions:** We recommend using a broad (ADRD) algorithm without CMS or medication data, particularly for epidemiological studies or a strict AD algorithm including CMS and medication cases for genetic discovery of late-onset AD associations in VA EMR, and a strict AD algorithm without CMS data for applications focused solely on AD and sensitive to misspecification. Careful evaluation of algorithm performance is warranted in different EMR systems, as ICD coding practices vary by institution, as demonstrated by this comparison of VA EMR and CMS data.

## Introduction

An estimated 7.2 million Americans age 65 and older have late-onset Alzheimer’s disease (AD)(1), the most common form of dementia. The prevalence of AD is expected to grow rapidly in the coming decades, with an estimated 13.8 million adults in the United States (US) developing AD by 2060(1). This increase will disproportionately impact the US Veteran population, as approximately half of the Veteran population is over the age of 65(2) and therefore at higher risk of AD and related dementias (ADRD). Moreover, relative to civilians, Veterans carry a higher burden of ADRD risk factors including depression, posttraumatic stress disorder (PTSD), traumatic brain injury (TBI), diabetes, and hypertension(3). ADRD in Veterans thus represents a significant public health burden, with substantial costs in medical care and caregiving within the US Department of Veterans Affairs (VA) healthcare system. Furthermore, as the largest integrated US healthcare system with an extensive electronic medical record (EMR) system, monitoring trends in ADRD rates and ADRD risk factors is key for VA planning and resource allocation and provides an important bellwether of the potential impact of ADRD in US healthcare more broadly.

Epidemiological studies of dementia often use International Classification of Diseases (ICD) codes to identify cases of dementia in EMR, both in studies based on the VA healthcare system and other systems(4–6). These codes are also used at the biobank level to perform phenome-wide association studies (PheWAS), which enhance insight into disease mechanisms by utilizing genetic data in conjunction with extensive EMR-derived phenotypes to detect broader health implications of genetic variation(7). Phenotype codes (PheCodes) are ICD-code based classifiers for a wide range of diseases developed with PheWAS as a use-case scenario(7). They have been extensively used in PheWAS studies(8, 9) as well as other studies of specific diseases(10, 11). Another typical use for ICD-code based algorithms is monitoring healthcare data for changes in disease rates over time and for policy planning. A specific example is the curated set of Chronic Conditions Warehouse (CCW) algorithms for identifying disease rates in Centers for Medicare & Medicaid (CMS) data. Although newer machine learning-based classifiers for identifying dementia are also being developed(12), the use of ICD-code based classification algorithms remains an important area of research.

Identifying ADRD in the VA EMR is particularly challenging. One study found widespread use of non-specific dementia codes (e.g., ICD-9 code 294.8: Other persistent mental disorders) and underuse of specific dementia diagnostic codes in the VA healthcare system(13). Another study found that a majority of patients with dementia in the VA healthcare system receive their initial diagnosis code from a primary care physician rather than a dementia specialist (e.g., neurologist, psychiatrist, or neuropsychologist), resulting in imprecise diagnoses (14).

Additionally, previously developed dementia algorithms have had a high false positive rate when used to identify dementia cases under age 60(4). Hence, when examining ICD codes for different dementias, we must view them as an imprecise tool and only probabilistically related to the true underlying pathology. Moreover, the probability of having a specific underlying form of dementia given the presence of an ICD code might differ based on the source of the code (e.g., general practice versus specialty clinics) and whether it is used to track treatment or for billing purposes.

In prior work, we developed novel ICD-code algorithms tailored for use in genetic studies of AD, ADRD, and dementias within the VA Million Veteran Program (MVP) and more generally in the VA healthcare system(15). We benchmarked our algorithms against several other pre-existing ICD-based methods and AD-medication-identified cases. In a large MVP cohort (N = ∼286,000), we performed a chart review of a subset of n = 103 participants to validate our MVP algorithms and assess their ability to detect apolipoprotein E (*APOE*) ε4 associations, the strongest genetic predictor of late-onset AD. Overall, we found that our newly developed AD and dementia algorithms were similar to, and sometimes outperformed, existing CCW and PheCode algorithms(13, 14). Specifically, our MVP-ADRD algorithm performed well in chart review and generated robust genetic signals, enhanced by the inclusion of medication-identified cases, and was in fact very comparable to the existing PheCode dementia algorithm. Performance of this algorithm was further improved when cases were restricted to onset age ≥60. We concluded that our algorithms would perform well in the broader VA and may also be suitable for other large-scale EMR-based studies of AD and dementia when biomarkers and imaging (e.g., amyloid positron emission tomography [PET] scans, cerebrospinal fluid [CSF]) are unavailable.

In this follow-up study, we extend our previous research and present new findings. We have updated our algorithms to reflect changes in ICD code usage (e.g., new ICD extensions which indicate severity; see Supplemental Table 1 for more detail on these changes); extended our chart review-based validation of our MVP AD and dementia algorithms; compared our MVP-derived algorithms to PheCode and CCW algorithms; and examined the impact of incorporating medication data into the algorithms. Moreover, we evaluated the effect of including CMS data as an additional source of EMR data, with the goal of preventing data gaps resulting from Veterans who receive their clinical care outside the VA healthcare system. Veterans who receive care outside VA may be systematically deidentified in VA-EMR-only studies, which may create a potential selection bias relevant to genetic and epidemiological studies. We expected CMS data would significantly increase our algorithms’ sensitivity and case yield, but whether CMS data, which includes ICD codes for Medicare/Medicaid reimbursement purposes, would have an impact on specificity remained an open question. Finally, we evaluated our algorithms by examining their association with pertinent AD genetic factors, including *APOE* ε4 and an AD polygenic risk score (PRS) that summarizes genetic risk due to other genetic variants, as an indirect indicator of the proportion of individuals with (true) late-onset AD in any particular (inferred) case set (to avoid confusion throughout, we will distinguish between those labeled as “cases” under any particular algorithm versus individuals who have a particular disease). As such, our work here specifically focuses on the algorithms’ utility for genetic studies of late-onset AD in VA but has broader implications for other EMR-based studies.

## Methods

### VA Million Veteran Program

MVP began enrolling participants in 2011 with the goal of establishing a mega-biobank to study how genes, lifestyle, military experiences, and exposures impact Veterans’ health and wellness(16). MVP is one of the largest and most genetically diverse biobanks in the world, and as of November 2023, had enrolled over one million Veterans, powering genetic studies on cancer, diabetes, PTSD, and a wide diversity of phenotypes (17). All MVP participants sign an informed consent document upon study enrollment. With almost half of the MVP cohort age 65 and older, and therefore at risk for developing AD and other dementias, MVP is a valuable resource for conducting studies on these diseases. This also points to the need for VA-specific AD and dementia algorithms that are attuned to the ways in which ICD codes are used across the VA healthcare system.

### Data Sources

#### VA EMR Data

The VA EMR dates back to the mid-1990s and is the primary source of phenotype data for MVP. After data access approval, MVP studies are provisioned VA-based data, including ICD-9/10 codes, treatment codes, and medication/pharmacy data from the EMR.

#### CMS Data

CMS data is also available and includes information on active MVP enrollees captured by Medicare or Medicaid, such as ICD code-based data from Medicare/Medicaid claims, Medicare/Medicaid-based prescription information, and demographics, beneficiary summaries, inpatient and outpatient visits, vital status, and facility and long-term care information(18). As of September 2021, just over half (52%) of Veterans Health Administration (VHA) enrollees were also enrolled in Medicare (48%) or Medicaid (4%)(19). The majority of older VHA enrollees are dually enrolled in VHA and Medicare; specifically, 91% of those ages 65-74 are and 94% of those ages 75-84 were dually enrolled(19). We used the following Medicare files to pull ICD-9/10 codes: (1) Fee-for-service (FFS) claims files: Inpatient, Skilled Nursing Facility (SNF), Home Health Agency (HHA), Hospice, Outpatient, Carrier, and Durable Medical Equipment (DME); and (2) Medicare Advantage (MA) encounters: Inpatient, SNF, HHA, Outpatient, Carrier, DME, and Part D Prescription Drug Events. Additionally, we used the following Medicaid files to pull ICD-9/10 codes: Other Services (OT), Long Term Care (LT), Inpatient (IP), and Prescription Drug (RX).

### AD and Dementia-Related Algorithms

We evaluated the following nine AD and dementia-related algorithms in this study: four MVP-derived algorithms, two CCW algorithms, two PheCode algorithms, and one AD medication-based algorithm.

#### MVP algorithms

Through our ongoing work in MVP(13, 20–23), we developed four AD-dementia phenotypes based on ICD codes for specific use within the VA EMR. These ICD code-based phenotypes are as follows, in order from most to least specific: (1) “MVP-AD”; (2) “MVP-AD+”, including AD and other non-specific dementia codes (e.g., dementia without behavioral disturbance); (3) “MVP-ADRD”, Alzheimer’s disease and related dementias; and (4) “MVP-Dementia.” See Tables 1-2 for a list of the ICD codes included in each phenotype. We required ≥2 ICD codes on different dates to qualify as a case at any of these four levels. Our four phenotypes are nested, such that anyone who is classified as an MVP-AD case will automatically also meet the criteria for MVP-AD+, MVP-ADRD, and MVP-dementia.

**Table 1.**
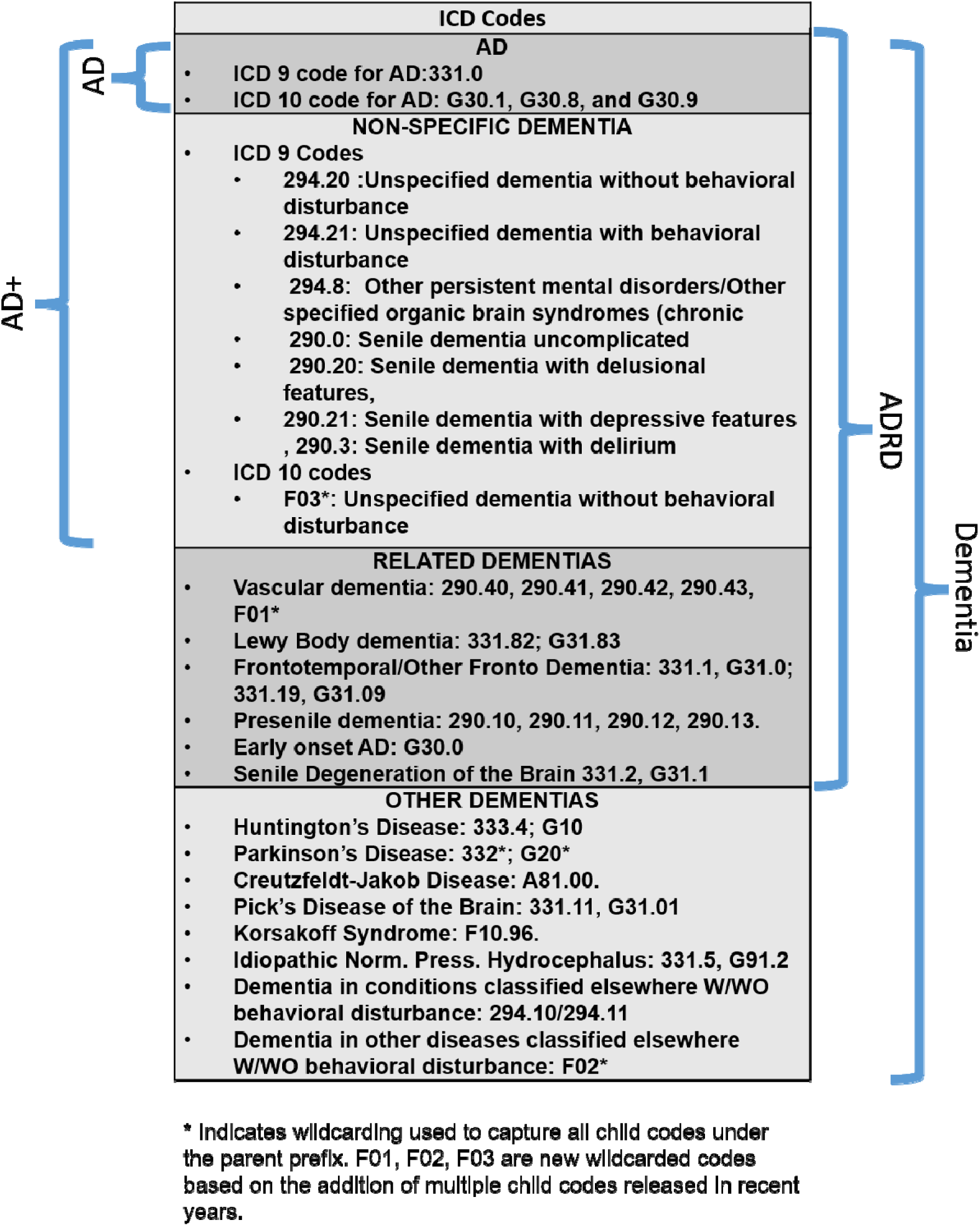
ICD-9/10 codes used in our derived algorithms for AD, AD+, ADRD, and dementia. Two or more ICD codes are required to qualify as a case in any of the nested categories.

**Table 2.**
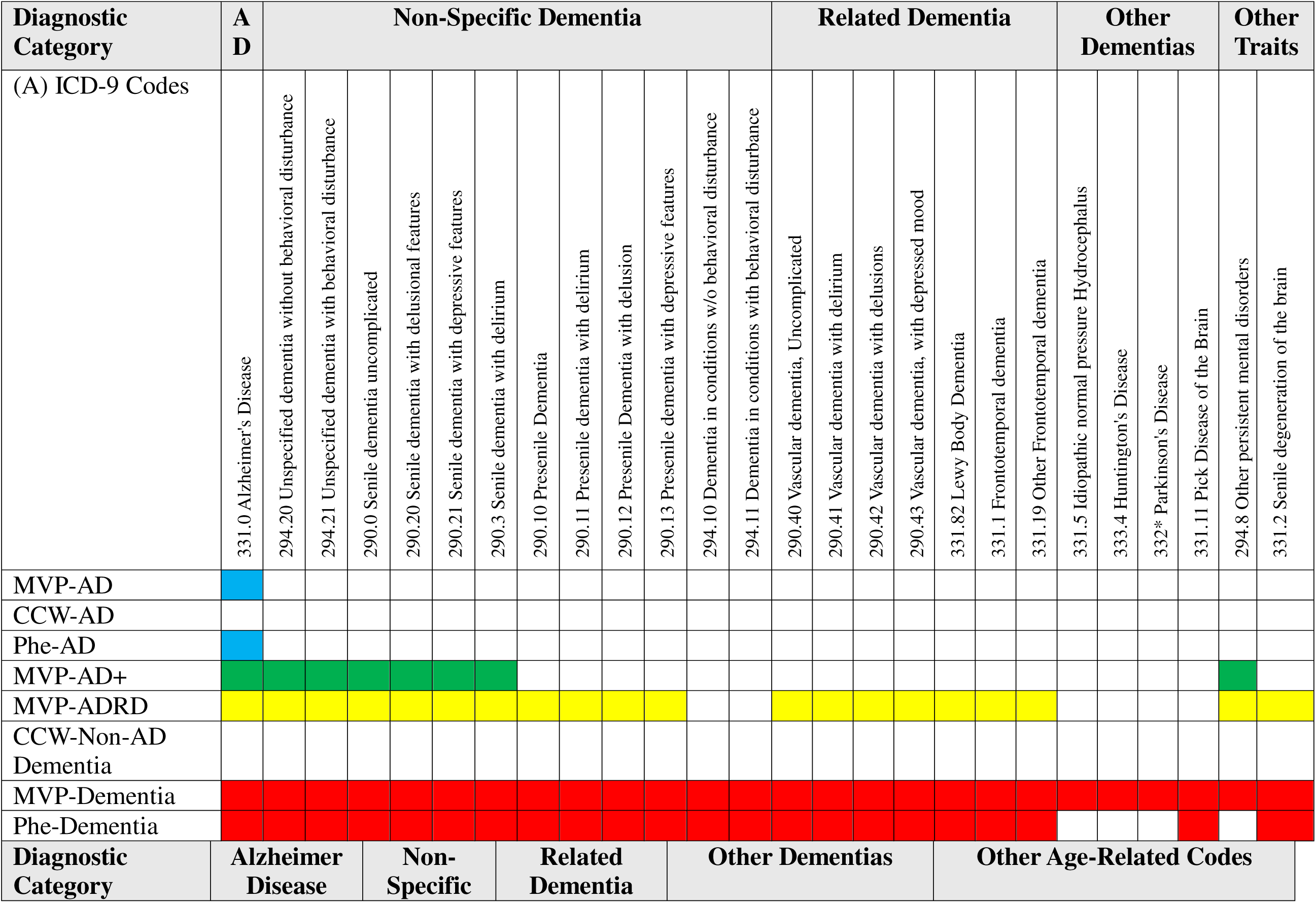
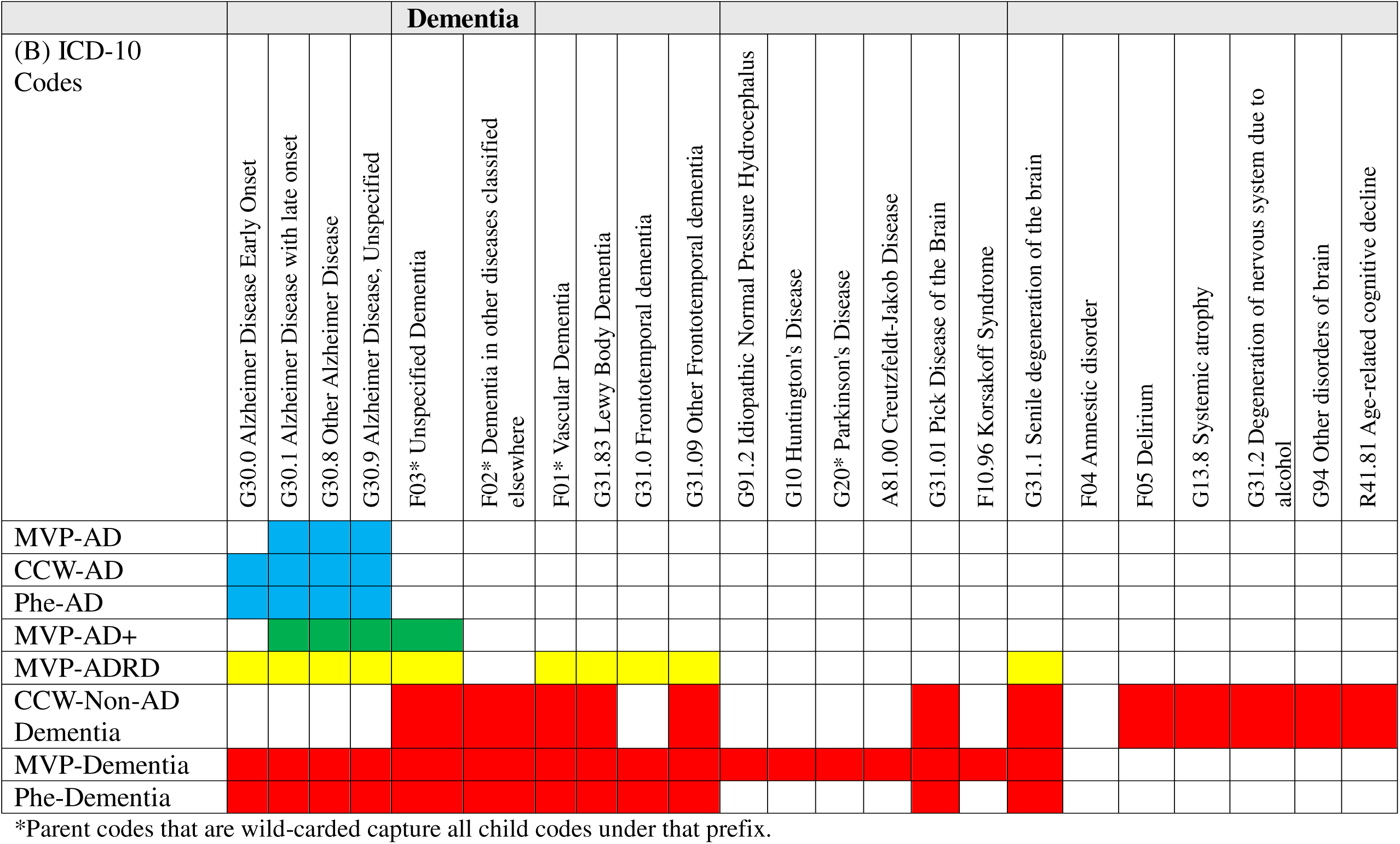
List of ICD codes included in the different classification algorithms including ICD-9 codes (A) and ICD-10 codes (B). Blue cells are ADspecific algorithms, green cells represent AD+, yellow cells represent ADRD algorithms, and red cells represent dementia algorithms.

#### CCW algorithms

The following AD and dementia-related algorithms from CCW were used, applying the most updated CCW definitions: (1) Alzheimer’s disease (hereafter referred to as “CCW-AD”; updated 02/2022) and (2) non-Alzheimer’s dementia (hereafter referred to as “CCW-Non-AD dementia”; updated 07/2023). Consistent with the updated CCW definitions, we required ≥1 inpatient ICD code or ≥2 outpatient ICD codes to be considered a case in the CCW algorithms. See Table 2 for a list of ICD codes included in each CCW algorithm (and Supplemental Table 1 for changes that were made to the CCW algorithms since our original chart review paper was published).

#### PheCode algorithms

The following PheCode algorithms were evaluated: (1) PheCode 290.11 (Alzheimer’s disease; hereafter referred to as “Phe-AD”) and (2) PheCode 290.1 (dementias; hereafter referred to as “Phe-Dementia”). We required ≥2 qualifying ICD codes on different dates to be considered a case in the PheCode algorithms. See Table 2 for a list of ICD codes included in each PheCode algorithm.

#### AD medication algorithm

We also identified AD medication cases (hereafter referred to as “AD-Med”) among MVP participants. This algorithm identified cases based on the prescription of an FDA-approved medication for AD treatment (at any time) in the VA EMR and/or CMS record, such as evidence of a prescription for a cholinesterase inhibitor (e.g., donepezil, galantamine, rivastigmine) or memantine. These AD-Med cases were not excluded from the ICD code cases.

To evaluate the performance of our MVP algorithms, we benchmarked our algorithms against the existing CCW and PheCode algorithms and AD-Med algorithm described above. Finally, we evaluated the performance of each algorithm with and without the inclusion of CMS data alongside VA EMR data (i.e., we used ICD codes from CMS data in combination with ICD codes from VA EMR data to update ICD code counts and onset dates to detect cases of AD/ADRD/Dementia in the cohorts).

### Chart Review

#### Cohort

The combined chart review cohort of n=203 included 100 newly reviewed participants (present study) and 103 participants from our original chart review study (Merritt et al., 2025) (15). The chart review cohort for the present study is based on the MVP v21_1 data release which includes MVP participants who enrolled prior to October 31, 2021 (n=840,866). This cohort comprises a random sample of Veterans with any of the ICD codes for AD/ADRD/Dementia as defined in our original chart-review project(15). See Supplemental Material Table 2 for a complete list of qualifying ICD-9/10 codes.

#### Procedures

Medical record notes, scrubbed of patient identifiers (names, birth dates, addresses), were provisioned to an MVP workspace for review. Only notes with relevant keywords related to AD and dementia were provisioned (see Supplemental Material Table 3 for details). Reviewers used a Microsoft Access template to view notes and document pertinent data from their review (see “Reviewer Template” in the Supplementary Material of (15)). The template allowed reviewers to note specific evidence of AD/dementia, such as the presence of a neuropsychological assessment, cognitive screening scores, brain imaging scan results, AD medications, and autopsy results. Using a “silver standard” classification system, each subject was classified as “Likely,” “Possible,” or “Likely Not” for both AD and all-cause dementia (see “Reviewer Guide” in the Supplemental Material for a detailed description of the guidelines and instructions reviewers used to classify AD and dementia cases). Two reviewers were assigned to each chart and shared their classifications at weekly consensus meetings. Initial disagreement was resolved through reviewer discussion until a consensus was reached. Inter-rater reliability (IRR) was calculated based on reviewers’ initial classifications, which included disagreements.

**Table 3.**
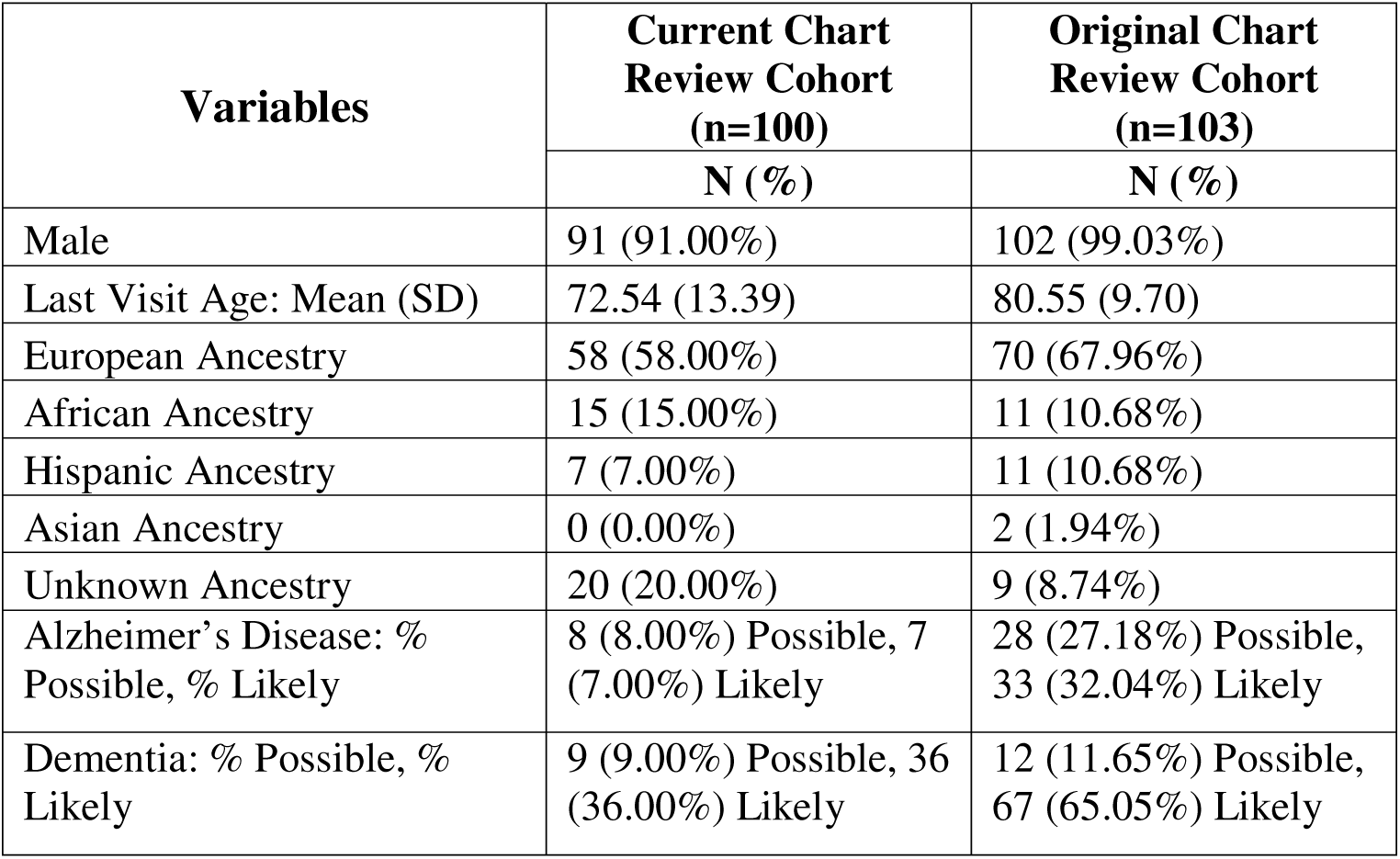
Demographics for MVP participants included in the current chart review project and the original chart review project (Merritt et al., 2025).

### MVP Genotype Data

All genetic data for this study were provisioned by the MVP Bioinformatics Core, who performed the data processing and data cleaning. See Hunter-Zinck et al. (2020) for details(24). Since the effect of *APOE* is known to differ by ancestry(25), we confined our genetic analyses to MVP participants of European ancestry, determined by the harmonized ancestry and race/ethnicity (HARE) method(26). We followed the same *APOE* genotyping and analysis process as described in detail in our original chart review project(15). Briefly, *APOE* genotypes were generated from SNP data extracted from the Phase 4 MVP genotype release (n = ∼650,000)(24). Imputation for the two SNPs used to determine the isoform (rs7412 and rs429358) was performed using the NHLBI Trans-Omics for Precision Medicine (TOPMed) reference panel(27), and are well imputed in MVP participants of European ancestry (imputation quality r^2^>.9)(20). *APOE* ε4 dosage (0, 1, or 2 alleles) was then coded for analysis. In addition to evaluating *APOE*, an AD polygenic risk score (PRS) was calculated based on the Kunkle et al. (2019) AD genome-wide association study (GWAS) (28), consistent with the methods used in our earlier studies(20). By excluding the *APOE* region on chromosome 19, this PRS summarizes the impact of other (non-*APOE*) known AD genetic risk loci. See our previous publications for more details on the generation of the AD PRS (20).

### Data Analyses

#### Chart Review Analyses

Our primary analysis classified participants as “Likely Not” AD or dementia to “Possible” and “Likely” AD or dementia following our established chart review framework [16]. As it is not reasonable to evaluate the performance of an AD-specific algorithm relative to a chart review for a broad dementia classification, and vice versa, we compared the AD algorithms (i.e., MVP-AD, CCW-AD, Phe-AD, and MVP-AD+) to the chart review-based diagnosis of AD, and the ADRD/dementia algorithms (i.e., MVP-ADRD, MVP-Dementia, CCW-Non-AD Dementia, Phe-Dementia) to the chart review-based dementia classification.

Additionally, we evaluated the performance of each algorithm based on VA data alone, or treatment codes from both VA and CMS data. The sensitivity, specificity, positive predictive value (PPV), negative predictive value (NPV), and IRR were calculated using R 4.4.1(29). IRR was calculated using the Cohen’s Kappa statistic, which incorporates multiple ordered levels of classification into the calculation (https://cran.r-project.org/web/packages/irr/index.html). In all cases, consensus was reached through discussion, and no charts required adjudication by a third reviewer.

#### Genetic Associations

We examined the strength of association of our MVP-derived algorithms and the previously derived PheCode and CCW algorithms with *APOE* ε4 and the AD PRS. All the different case sets according to the various algorithms were compared to a common control set. Using a logistic regression model, *APOE* ε4 allele dosage was first tested in association with dementia case/control status, using *APOE* ε4 allele dosage as a continuous predictor, age as a covariate, and the various AD and dementia algorithms as the outcome variable versus controls. Additional analyses examined the impact of including CMS data, including an age restriction for cases (first dementia ICD code at age 60+ or 65+), and combining ICD code cases of AD, ADRD, and dementia together with AD-Med cases. When examining the impact of age thresholds and including CMS data, we adjusted the control sets to match these new conditions. That is, we required that the controls’ last reported VA visit also exceed the age threshold for the thresholding analyses, and we also excluded controls who had a qualifying ICD code in either the VA-EMR or CMS when compared to cases identified using CMS data. Finally, to ensure that our comparisons did not apply to *APOE* alone, we examined associations between an AD PRS and the various AD and dementia algorithms using an identical progression of analyses.

## Results

### Cohort

The majority of participants (91%) in the current cohort were male, and the mean age at last recorded visit was approximately 73 years (Table 3). Over half of the participants were of European ancestry (58%), followed by African ancestry (15%) and Hispanic ancestry (7%); 20% were not classified into any of the specific ancestry groups. In this cohort, 8% of participants were classified as “Possible” AD, 7% as “Likely” AD, 9% as “Possible” Dementia, and 36% as “Likely” Dementia. These rates are lower than those observed in the chart review cohort for our original paper, which used different methods for choosing individuals to review, prioritizing those with a moderate level of evidence for AD or dementia(15).

### Chart Review Results

The IRR was high for both AD (kappa = 0.78) and dementia (kappa = 0.84). The frequencies of algorithm classifications (Table 4) were examined within the current chart review cohort (n=100), our original chart review cohort (n=103), the overall MVP v24_1 participant cohort (n=1,010,283), and an age-restricted cohort of MVP participants age 65+ (n=656,767). When considering the overall MVP cohort, the CCW-AD algorithm (in VA EMR data and VA EMR/CMS data) identified the largest number of AD cases (n=25,542). For ADRD/dementia, our MVP-Dementia algorithm (in VA EMR and VA EMR/CMS data) identified the largest number of cases (n=71,621 for VA EMR, n=96,037 for VA EMR/CMS). The addition of CMS data had the largest impact on the AD algorithms, either doubling or nearly doubling the number of identified cases for the MVP-AD (from 11,345 to 22,537), CCW-AD (from 13,314 to 25,542), and Phe-AD algorithms (from 11,591 to 23,101). The addition of CMS data had the smallest impact on the AD-Med algorithm, adding only 317 cases (see Table 4), as the amount of prescription data in CMS was low relative to the VA data.

**Table 4.** Prevalence of AD and Dementia as classified by each algorithm compared to the chart review diagnoses in the Current Chart Review Cohort, the Original Chart Review Cohort, the entire MVP cohort, and a cohort of MVP participants age 65 and older.

| **Algorithm** | **Current**<br>**Chart**<br>**Review**<br>**Cohort**<br>**(n=100)**<br>**N (%)** | **Original**<br>**Chart**<br>**Review**<br>**Cohort**<br>**(n=103)**<br>**N (%)** | **MVP Cohort**<br>**(n=1,010,283)**<br>**N (%)** | **MVP Cohort**<br>**Age ≥ 65**<br>**(n=656,767)**<br>**N (%)** |
| --- | --- | --- | --- | --- |
| MVP-AD (VA EMR) | 11 (11.00) | 49 (47.57) | 11,345 (1.12) | 11,201 (1.71) |
| MVP-AD (VA EMR/CMS) | 16 (16.00) | 62 (60.19) | 22,537 (2.23) | 22,298 (3.40) |
| CCW-AD (VA EMR) | 11 (11.00) | 60 (58.25) | 13,314 (1.32) | 13,070 (1.99) |
| CCW-AD (VA EMR/CMS) | 17 (17.00) | 70 (67.96) | 25,542 (2.53) | 25,171 (3.83) |
| Phe-AD (VA EMR) | 11 (11.00) | 49 (47.57) | 11,591 (1.15) | 11,409 (1.74) |
| Phe-AD (VA EMR/CMS) | 16 (16.00) | 63 (61.17) | 23,101 (2.29) | 22,814 (3.47) |
| MVP-AD+ (VA EMR) | 41 (41.00) | 80 (77.67) | 44,573 (4.41) | 42,527 (6.48) |
| MVP-AD+ (VA EMR/CMS) | 47 (47.00) | 84 (81.55) | 68,439 (6.77) | 65,918 (10.04) |
| MVP-ADRD (VA EMR) | 46 (46.00) | 83 (80.58) | 52,536 (5.20) | 49,887 (7.60) |
| MVP-ADRD (VA EMR/CMS) | 51 (51.00) | 86 (83.50) | 77,030 (7.62) | 73,813 (11.24) |
| MVP-Dementia (VA EMR) | 58 (58.00) | 85 (82.52) | 71,621 (7.09) | 67,182 (10.23) |
| MVP-Dementia (VA EMR/CMS) | 62 (62.00) | 89 (86.41) | 96,037 (9.51) | 90,892 (13.84) |
| CCW-Non-AD Dementia (VA EMR) | 39 (39.00) | 68 (66.02) | 57,356 (5.68) | 54,272 (8.26) |
| CCW-Non-AD Dementia (VA EMR/CMS) | 44 (44.00) | 76 (73.79) | 88,760 (8.79) | 84,768 (12.91) |
| Phe-Dementia (VA EMR) | 45 (45.00) | 84 (81.55) | 52,063 (5.15) | 49,550 (7.54) |
| Phe-Dementia (VA EMR/CMS) | 50 (50.00) | 93 (90.29) | 77,817 (7.70) | 74,678 (11.37) |
| AD-Med (VA EMR) | 31 (31.00) | 64 (62.14) | 41,306 (4.09) | 38,699 (5.89) |
| AD-Med (VA EMR/CMS) | 31 (31.00) | 64 (62.14) | 41,623 (4.12) | 38,931 (5.93) |

The sensitivity, specificity, PPV, and NPV of the various AD and dementia algorithms relative to chart review classifications of “Likely Not” versus “Possible/Likely” AD and dementia are presented in Figure 1A-1D and Supplemental Material Table 6. Panels A and B of Figure 1 present the sensitivity and 1-specificity for comparison with AD chart review classifications and dementia chart review classifications, respectively. Panels C and D of Figure 1 represent the PPV and NPV for algorithms compared to the chart review classifications of AD and dementia. As before(15), the use of AD medications by itself had poor performance relative to other algorithms. That is, it had lower specificity and NPV than algorithms with similar sensitivity and PPV, respectively. Adding AD-Med cases to the ICD code algorithms (red arrows) and incorporating CMS data (blue arrows) with the ICD code algorithms resulted in increased sensitivity and NPV, but with trade-offs between specificity and PPV.

**Figure 1.**
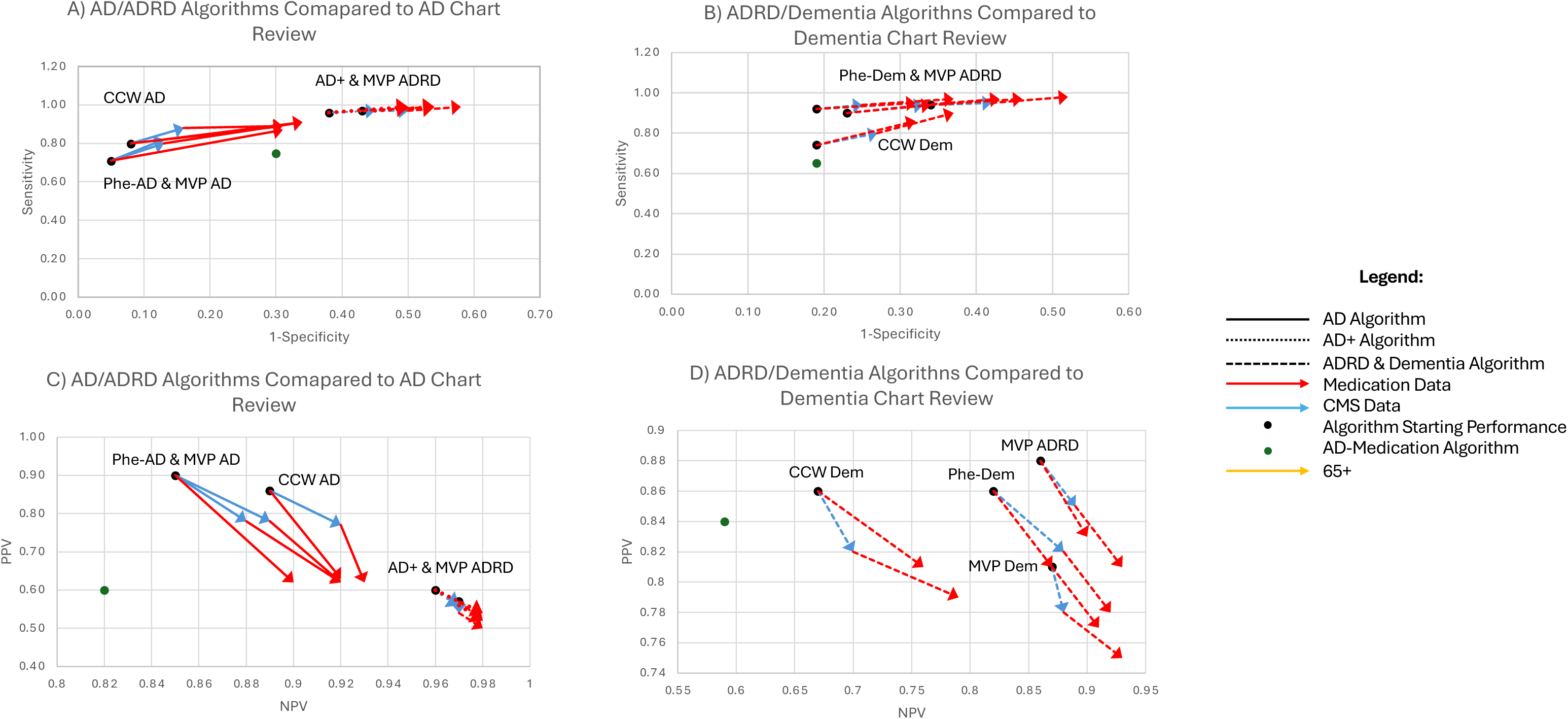
Chart review results and comparison of algorithm performance in classifying AD (A and C) and dementia (B and D). Sensitivity versus 1-Specificity are presented in A and B, and positive predictive value (PPV) and negative predictive value (NPV) are presented in C and D. The starting points for algorithm performance are marked with black dots; the end point of each arrow represents the final algorithm performance after the addition of either medication or CMS data. The impact of including medication cases is presented in red arrows, and the impact of adding CMS data to VA EMR data is presented in blue arrows. The performance of using AD medication prescription information alone to identify cases is presented with a green dot.

When examining the performance of the algorithms in identifying AD cases (Figure 1A and 1C, Supplemental Material Table 6), the addition of medication cases and CMS data had a larger impact on the performance of the AD algorithms than the AD+ and ADRD algorithms. The performance of Phe-AD and MVP-AD were very similar. CCW-AD, which allows for cases with a single inpatient ICD code and also excludes ICD-9 codes, had slightly higher sensitivity than the other two algorithms, lower specificity, slightly lower PPV, and lower NPV.

When compared to the chart review classifications of dementia (Figure 1B and 1D, Supplemental Material Table 6), the MVP-Dementia and Phe-Dementia algorithms had relatively similar performance, both with and without the inclusion of CMS data and medication data, with the MVP-ADRD algorithm having somewhat better performance than the Phe-Dementia algorithm, especially in terms of PPV and NPV. The MVP-ADRD algorithm performed better than the CCW-Non-AD Dementia algorithm, which had similar specificity and PPV to Phe-Dementia, but lower sensitivity and NPV, likely due to the exclusion of AD codes.

### Genetic Results

The significance of the associations between *APOE* ε4 and the diagnostic algorithms for MVP participants of European ancestry is presented in Figure 2A and Supplementary Material Table 4. The case definitions for each algorithm were strongly associated with *APOE* ε4 (all p<10^-300^), and all associations exceeded the maximum precision of standard floating-point computation, so Z-scores were used to compare the relative strength of association. Based on the VA EMR data only (i.e., prior to the addition of CMS data, the age cutoff, and combining the medication and ICD-code based cases), the Phe-Dementia and MVP-ADRD algorithms produced the strongest association with *APOE* ε4 (Z scores of 54.79 and 54.04, respectively). In contrast, the MVP-AD and Phe-AD algorithms produced the weakest association with *APOE* ε4, but with the largest observed ORs (MVP-AD: Z=44.86, OR=2.40[2.31,2.50]; Phe-AD: Z=45.42, OR=2.41[2.32,2.50]). The AD-Med algorithm also showed strong associations with *APOE* ε4 (Z=52.11). The broader dementia algorithms (MVP-Dementia and CCW-Non-AD Dementia) had a similar strength of association as the stricter AD algorithms, but with much lower ORs (MVP-Dementia: OR=1.54[1.52,1.57]; CCW-Non-AD Dementia: OR=1.71[1.68,1.74]; MVP-AD: OR=2.40[2.31,2.50]; Phe-AD: OR=2.41[2.32,2.50]).

**Figure 2.**
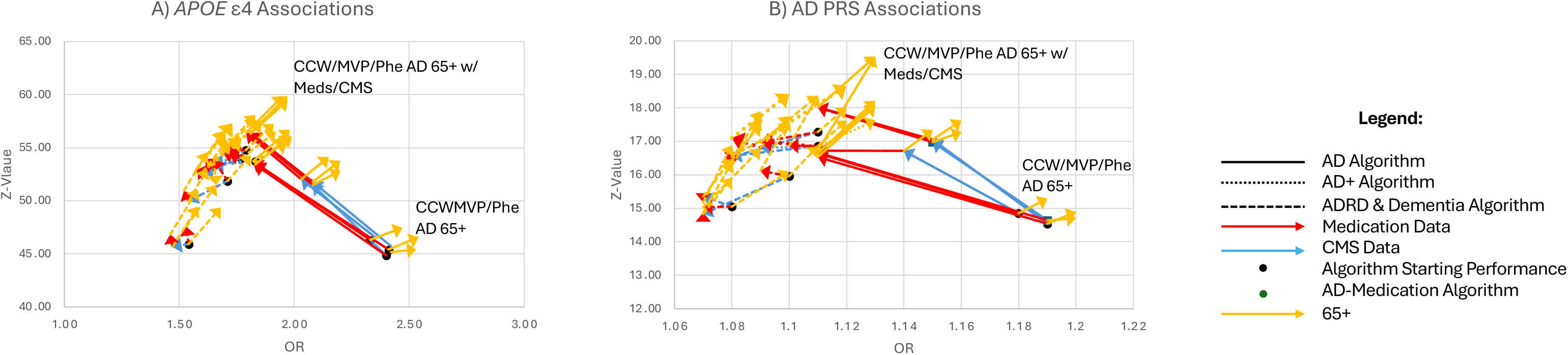
Association between algorithm-defined case status and (A) *APOE* ε4 and (B) an AD PRS for the various AD and dementia algorithms. The starting points for algorithm performance are marked with black dots; the end point of each arrow represents the final algorithm performance after the addition of either medication or CMS data or restricting the sample to age 65+. The impact of including medication cases is presented in red arrows, the impact of adding CMS data to VA EMR data is presented in blue arrows, and the impact of restricting cases to be age 65+ is presented in yellow arrows.

The addition of medication cases to the ICD code algorithms and the addition of CMS data increased the case numbers substantially but both were associated with lower ORs for the *APOE* and AD PRS analyses (Figure 2A and 2B, Supplementary Material Table 4 & 5). However, the impact on the strength of genetic association differed somewhat, as both increased the strength of association substantially for the strict AD algorithms (MVP-AD, CCW-AD, and Phe-AD) but had a relatively smaller effect on the broader dementia definitions, with slight increases observed due to the addition of the AD medication cases and slight decreases due to the addition of CMS data to most of the broader ICD code algorithms. The use of an age cutoff of 60 or 65+ uniformly increased significance of the association with *APOE* ε4 and increased the OR estimates. The increase due to imposing an age cutoff was very similar whether the cutoff was 60+ or 65+, hence we have only plotted 65+ in Figure 2.

Which algorithms performed the best in terms of strongest genetic associations differed somewhat from our previous paper. Before, we observed the strongest association with the combination of MVP-ADRD and Phe-Dementia with AD medications and a 65+ age cutoff. Here, the addition of CMS data increased the sample sizes of the strict AD algorithms to where they could compete with the broader MVP-ADRD and Phe-Dementia algorithms. The three strict AD algorithms using VA EMR and CMS data supplemented by AD medications with an age 65+ cutoff performed very similarly to each other and produced the strongest genetic associations (*APOE* ORs from 1.95[1.91,2.00] for CCW-AD to 1.97[1.93,2.01] for Phe-AD and MVP-AD; Zs from 59.45 to 59.87 for MVP-AD and CCW-AD, respectively.) However, the observed ORs were still much lower than the strict AD algorithms with the 65+ cutoff without any CMS or medication data (*APOE* ORs from 2.47[2.38,2.56] for CCW-AD to 2.54[2.45,2.65] for Phe-AD. The previous best-performing algorithms in terms of greatest significance, MVP-ADRD and Phe-Dementia algorithms without CMS data (with medications and an age cutoff), had similar performance to the AD algorithms with CMS data (*APOE* OR=1.83[1.80,1.87] for both MVP-ADRD and Phe-Dementia; Zs from 57.75 for MVP-ADRD to 58.10 for Phe-Dementia), and lower performance if the CMS data is included (*APOE* OR=1.70[1.67,1.73] for MVP-ADRD and OR=1.69[1.66,1.71] for Phe-Dementia; Zs from 56.60 to 56.50, for MVP-ADRD and Phe-Dementia, respectively).

In the PRS analyses, the highest performing algorithm in terms of strength of association was again the three strict AD algorithms using VA EMR and CMS data supplemented by AD medications with an age 65+ cutoff (PRS OR=1.13[1.12,1.14] for CCW-AD, OR=1.13[1.12,1.15] for both MVP-AD and Phe-AD; Zs from 19.53 for MVP-AD and Phe-AD to 19.63 for CCW-AD). The second highest association was with the AD+ algorithm with medications and an age cutoff (PRS OR=1.10[1.09,1.11]; Z= 18.41), while the MVP-ADRD and Phe-Dementia with medications and an age cutoff fell to third and fourth place, again with very similar performance to each other (PRS OR=1.09[1.08,1.10] for both; Zs from 17.64 for Phe-Dementia to 17.89 for MVP-ADRD). In PRS association analyses without age cutoffs, the highest ORs were again observed for the strict AD algorithms, without medication and without CMS data (PRS ORs=1.19[1.16,1.22] for MVP-AD and Phe-AD, OR=1.18[1.16,1.21] for CCW-AD.

## Discussion

Dementia classification systems based on medical records and treatment codes remain relevant and important for the purposes of large-scale genetic and epidemiological studies and for monitoring the EMR for changes in diagnostic trends or rates of dementia. In this study, we have increased the sample size of our chart review cohort and increased the number of cases for genetic validation (i.e., from n= 405,540 MVP participants age ≥ 65 in our original chart review project to n= 656,767 in the current chart review project). When evaluating the frequency of AD and dementia cases as classified by each algorithm compared to the chart review diagnoses, we showed that AD is under-coded relative to non-specific dementia in the VA EMR, as the sensitivity increased from 0.71 for MVP-AD to 0.97 for MVP-ADRD, further supporting previous findings(13, 14). This issue of non-specific coding is not unique to the VA healthcare system and points to the importance of using more broadly defined algorithms, like our MVP-ADRD algorithm, to capture more cases for use in epidemiological studies of ADRD.

In this updated effort, we were specifically interested in determining whether adding CMS data would improve the performance of our algorithms, especially given the large increases in the number of cases that accompanied the inclusion of CMS treatment codes as input. Use of CMS data did indeed induce large changes in the number of cases identified across the different algorithms. We also found that adding CMS data displayed the classic tradeoff between sensitivity and specificity, similar to, but perhaps not as striking as, the addition of medication cases to the existing algorithms. Notably, this trade-off was not inevitable; if the lower than expected number of cases reflected under-coding of true AD, then we might expect CMS data to increase sensitivity with little to no decrease in specificity. This is clearly not the case. Similarly, if the increase in strict AD cases due to the addition of CMS data simply represented an increase in sample size for a similar new set of cases, that is, if the CMS-identified cases had the same underlying true rates of late onset AD, we would expect a larger boost in significance for the *APOE* ε4 and AD case associations. Instead, we observed a somewhat more modest increase in significance and a corresponding decrease in the estimated OR for AD. The results of the chart review and the *APOE* associations are most consistent with an increase in both the number of cases and a decrease in the likelihood of those being classified as cases having late-onset AD. One possibility is that the effect is due to some other difference between the AD cases identified using VA EMR data and those identified with VA EMR/CMS. However, both sets of cases are genetically confirmed European-ancestry Veterans, and the same pattern is seen both with the *APOE* ε4 associations and the AD PRS associations. The sets of cases overlap, so that many people who are identified as cases in CMS would also be identified as cases in the VA EMR, and many of the records included in CMS actually represent care provided within the VA healthcare system and reported through CMS for those covered by Medicare/Medicaid.

It is also worth examining the addition of medication and CMS data on the ADRD and dementia algorithms, to contrast with the way they impacted the stricter AD algorithms. The ADRD and dementia algorithms, which had larger case numbers to begin with because of their broader inclusion criteria, had smaller changes in sample size, *APOE* OR, and *APOE*-association significance after the addition of medication and CMS data. That is, in some sense, they were already further ahead on a similar trajectory, so that the inclusion of medication and CMS data was unnecessary. This, along with the lower ORs observed for the strict AD cases, leads us to conclude that the inclusion of CMS data and medications to the stricter AD criteria represents a broadening of the criteria rather than simply an increase in the number of AD cases. Indeed, the ADRD and dementia algorithms and the stricter AD algorithms appear to be converging to a similar peak in Figure 2A and 2B. This has several implications above and beyond which algorithms perform the best. First, following on a theme we noted in our previous paper, the composition of the case group does not simply depend on the category of included ICD codes. When we add CMS data to the strict AD algorithms, they become more ADRD-like, detecting more cases but with a smaller proportion of individuals with late-onset AD, as indicated by the *APOE* ε4 OR. So, based on this, we find that the broader dementia categories with CMS data have very similar performance to the stricter AD algorithms with the CMS data, and, depending on the situation, one or the other could be used. However, the higher ORs noted for AD algorithms based only on the VA EHR indicate that those algorithms should only be used with VA data if the particular study is sensitive to the proportion of other dementias being included in the case set. Regardless of the exact reason for the lower specificity when CMS data is added to AD algorithms or the reduced OR in *APOE* associations, the implications are largely the same: the increase in sample sizes due to the inclusion of CMS data cannot be simply viewed the same as increasing the sample sizes for the cohort under investigation. The addition of CMS also likely changes the underlying mix of dementias included in ADRD algorithms and the proportion of individuals with late-onset AD in those identified as cases using the stricter AD algorithms. Determining which algorithm is most appropriate for a specific purpose requires not only choosing a category of included codes but also evaluating how these codes perform and how they are being applied in practice, which varies across different medical systems and data sources.

It is possible that the relative difference in the ORs and lower specificity we observed when using CMS data is due to differences in CMS coding and reimbursement of AD versus other forms of dementia(30). Other studies have found that individuals with less severe AD are less likely to be recorded as having AD in CMS data, and that CMS data has low specificity for differentiating dementia versus mild cognitive impairment (MCI)(31). Finally, CMS coding practices may vary regionally, across different care settings, and by different specialists, leading to inconsistencies that impact the association of case algorithms with *APOE* ε4.

While our study has many strengths, we must also note the limitations. Using ICD-code based algorithms and AD medication data to identify AD and dementia cases and controls utilizes only one part of the myriad information contained in the EMR, distinct from machine learning algorithms or natural language processing methods that identify and/or predict dementia cases based on a more thorough data extraction, including medical/provider notes and other unstructured data(32, 33). However, because ICD-code algorithms continue to be used in epidemiological studies(4, 34), PheWAS, and disease monitoring and planning(35), we believe that the continued refinement and validation of ICD-code algorithms for ADRD remains important.

Conducting our chart review project entirely within the MVP cohort also holds inherent limitations because MVP participants are not a random sample of VHA users, which may restrict the participants available for inclusion in the chart review. Another limitation lies in the fact that genetic associations were only evaluated for European-ancestry participants, potentially limiting the generalizability of these results to non-European individuals. Our chart review incorporated many different AD identification factors, including neuropsychological assessments, neuroimaging scans, cognitive screening assessments, AD medication prescription history, and autopsy results. However, these factors were not uniformly available across all participants, making our chart review less robust than other studies in which standardized neuropsychological testing, clinical examination, *APOE* genotyping, and other data are available for all participants(36).

## Conclusions

In the present study, we continued to refine and validate our MVP algorithms for AD and dementia in MVP and analyzed their associations with genetic risk for AD. Mindful of the fact that a significant portion of the Veteran population is dually enrolled in Medicare/Medicaid, we extended our work to evaluate the performance of our MVP algorithms with the addition of CMS data. Supplementing these diagnostic algorithms with CMS data led to increased sample sizes and higher sensitivity, but at the cost of lower specificity. Additionally, we observed that CMS data weakened the strength of genetic associations for most individual algorithms for ADRD and dementia but bolstered the associations for the strict AD algorithms (i.e., MVP-AD, AD-Med/MVP-AD, CCW-AD and Phe-AD). Taken together, we conclude that the use of CMS data in MVP is not strictly necessary, especially for genetic studies, particularly with ADRD. Moving forward, our recommendation is to use our MVP-AD algorithm with CMS and medication data for genetic discovery in VA/MVP-based epidemiological studies of AD and ADRD if CMS data is available. If CMS data is not available, we recommend using MVP-ADRD or Phe-Dementia.

While genetic validation is most informative for genetic studies, we recommend using ADRD (VA EMR alone) and/or AD algorithms with CMS for epidemiological studies, depending on whether the study design prioritizes sensitivity versus specificity. The choice as to which AD algorithm is used makes little difference (MVP, PheCode, or CCW). Regardless of the algorithm choice, the inclusion of CMS data results in a more ADRD-like phenotype, and an age cut-off should be applied in all late onset ADRD studies.

Altogether, the results of this chart review project support the continued use of broader ADRD classification algorithms when engaged in epidemiological studies using VA EMR data due to the persistent under-coding of AD. We also encourage the requirement of using more than one ICD code to qualify as a case; using AD medication to identify additional cases; and implementing age cutoffs for dementia onset. These algorithm best practices (i.e., requiring multiple ICD codes, integrating medication data, and applying age cutoffs) offer a validated and practical framework for identifying dementia cases in VA EMR research.

## Conflicts of Interest

The author(s) declared no potential conflicts of interest with respect to the research, authorship, and/or publication of this article.

## Data Availability

The data and code used to generate MVP results are accessible to researchers with MVP data access. Due to VA policy, MVP is currently only accessible to researchers with a funded MVP project (e.g., VA Merit Award, Career Development Award, NIH R01). Thus, the datasets generated and/or analyzed during the current study are not publicly available, but the corresponding author is willing to engage with reasonable requests, share code, and answer questions about the present study. Information about accessing MVP data can be found at: https://www.mvp.va.gov/pwa/joinmvp.

## Funding

This research is based on data from the Million Veteran Program, Office of Research and Development, Veterans Health Administration, and was supported by MVP000, as well as award I01BX005749 (MVP040) and NIH U24AG074855. We thank the Veterans who generously agreed to participate in the Million Veteran Program. This publication does not represent the views of the Department of Veterans Affairs or the United States Government.

Support for CMS and USRDS Data provided by the Department of Veterans Affairs, Veterans Health Administration, Office of Research and Development, VA Information Resource Center (Project Numbers SDR 02-237 and 98-004).

## Disclosures

JAL reports grants from Alnylam Pharmaceuticals, Inc., AstraZeneca Pharmaceuticals LP, Biodesix, Inc, Janssen Pharmaceuticals, Inc., Novartis International AG, Parexel International Corporation through the University of Utah or Western Institute for Veteran Research outside the submitted work.

DM is the co-founder of Remy Care, LLC.

## Acknowledgements

**MVP Cognitive Decline and Dementia during Aging Working Group Members:** Mark W. Logue Chair, Richard L. Hauger Co-Chair, Matthew Panizzon Secretary, Victoria Merritt Founding Member.

Members: Karl Brown, Catherine Chanfreau, Kelly Cho, Royce Clifford, Lindsay Farrer, Jennifer Fonda, Margaret Gillis, Kelly Harrington, Yuk-Lam Ho, William Kremen, Sophia Lee, Lauren Loeffel, Francesca Lopez, Julie Lynch, Adam Maihofer, David Marra, Jesse Mez, Mark Miller, Zoe Neale, Caroline Nievergelt, David Salat, Richard Sherva, Debby Tsuang, Meghan Wilkinson, Erika Wolf, Rui Zhang, Qing Zeng.

## VA Million Veteran Program

### Core Acknowledgements

#### MVP Program Office

- Sumitra Muralidhar, Ph.D., Program Director

US Department of Veterans Affairs, 810 Vermont Avenue NW, Washington, DC 20420

- Jennifer Moser, Ph.D., Associate Director, Scientific Programs

US Department of Veterans Affairs, 810 Vermont Avenue NW, Washington, DC 20420

- Jennifer E. Deen, B.S., Associate Director, Cohort & Public Relations

US Department of Veterans Affairs, 810 Vermont Avenue NW, Washington, DC 20420

#### MVP Steering Committee

- Co-Chair: Philip S. Tsao, Ph.D.

VA Palo Alto Health Care System, 3801 Miranda Avenue, Palo Alto, CA 94304

- Co-Chair: Sumitra Muralidhar, Ph.D.

US Department of Veterans Affairs, 810 Vermont Avenue NW, Washington, DC 20420

- J. Michael Gaziano, M.D., M.P.H.

VA Boston Healthcare System, 150 S. Huntington Avenue, Boston, MA 02130

- Adriana Hung, M.D., M.P.H.,

VA Tennessee Valley Healthcare System, 1310 24th Avenue, South Nashville, TN 37212

- Dave Oslin, M.D.

Philadelphia VA Medical Center, 3900 Woodland Avenue, Philadelphia, PA 19104

- Deepak Voora, M.D.

Durham VA Medical Center, 508 Fulton Street, Durham, NC 27705

#### MVP Co-Principal Investigators

- J. Michael Gaziano, M.D., M.P.H. VA Boston Healthcare System, 150 S. Huntington Avenue, Boston, MA 02130

- Philip S. Tsao, Ph.D. VA Palo Alto Health Care System, 3801 Miranda Avenue, Palo Alto, CA 94304

#### MVP Core Operations

- - Jessica V. Brewer, M.P.H., Director, MVP Cohort Operations VA Boston Healthcare System, 150 S. Huntington Avenue, Boston, MA 02130

- - Mary T. Brophy M.D., M.P.H., Director, VA Central Biorepository VA Boston Healthcare System, 150 S. Huntington Avenue, Boston, MA 02130

- - Kelly Cho, M.P.H, Ph.D., Director, MVP Phenomics VA Boston Healthcare System, 150 S. Huntington Avenue, Boston, MA 02130

- - Lori Churby, B.S., Director, MVP Regulatory Affairs VA Palo Alto Health Care System, 3801 Miranda Avenue, Palo Alto, CA 94304

- - Jacob T. Kean, Ph.D., Acting Director, VA Informatics and Computing Infrastructure (VINCI)VA Salt Lake City Health Care System, 500 Foothill Drive, Salt Lake City, UT 84148

- - Saiju Pyarajan Ph.D., Director, Data and Computational Sciences VA Boston Healthcare System, 150 S. Huntington Avenue, Boston, MA 02130

- - Robert Ringer, Pharm.D., Director, VA Albuquerque Central Biorepository New Mexico VA Health Care System, 1501 San Pedro Drive SE, Albuquerque, NM 87108

- - Luis E. Selva, Ph.D., Director, MVP Biorepository Coordination VA Boston Healthcare System, 150 S. Huntington Avenue, Boston, MA 02130 Shahpoor (Alex) Shayan, M.S., Director, MVP PRE Informatics VA Boston Healthcare System, 150 S. Huntington Avenue, Boston, MA 02130

- - Brady Stephens, M.S., Principal Investigator, MVP Information Center Canandaigua VA Medical Center, 400 Fort Hill Avenue, Canandaigua, NY 14424

- - Stacey B. Whitbourne, Ph.D., Director, MVP Cohort Development and Management VA Boston Healthcare System, 150 S. Huntington Avenue, Boston, MA 02130

## Notes

### Competing Interest Statement

The authors have declared no competing interest.

### Author Declarations

This study was approved by VA Boston Healthcare Research and Development Committee and US Department of Veterans Affairs Central IRB.

