## Supplementary File for "Chart review and genetic validation of electronic medical record dementia diagnoses in VA: The impact of CMS data"

### Supplemental Material

**Supplemental Table 1.** Changes in Chronic Conditions Warehouse (CCW) Algorithms

**Supplemental Table 2.** ICD-9/10 Codes Used in Original Chart Review Project

**Supplemental Table 3.** List of Concept Unique Identifiers (CUIs)

**Supplemental Table 4.** Algorithm Performance in MVP Participants of European Ancestry and *APOE* ε4 Associations

**Supplemental Table 5.** Algorithm Performance in MVP Participants of European Ancestry and AD Polygenic Risk Score (PRS) Associations

**Supplemental Table 6.** Algorithm Performance in Chart Review Cohort

**Reviewer Guide**

**Supplemental Table 1.** Comparison of algorithms in and CC30*. The discontinued Alzheimer's Disease and Related Disorders or Senile Dementia (CC27) classification is compared to the Non-Alzheimer's Dementia (CC30) new classification category. Italicized ICD codes represent codes that were present in both CC27 and CC30 algorithms; bolded ICD codes represent codes that are distinct between CC27 and CC30.

| **Algorithm** | **CC27 (1991-2021)** | **CC30 (2017-present)** | **Description of Changes** |
|---|---|---|---|
| Alzheimer's Disease | **331.0**, *G30.0, G30.1, G30.8, G30.9* (any DX on the claim) | *G30.0, G30.1, G30.8, G30.9* (any DX on the claim) | Removed 1 ICD-9 code |
| Alzheimer's Disease and Related Disorders or Senile Dementia (CC27) / Non-Alzheimer's Dementia (CC30) | **331.0, 331.11, 331.19, 331.2, 331.7, 290.0, 290.10, 290.11, 290.12, 290.13, 290.20, 290.21, 290.3, 290.40, 290.41, 290.42, 290.43, 294.0, 294.10, 294.11, 294.20, 294.21, 294.8, 797,** *F01.50, F01.51, F02.80, F02.81, F03.90, F03.91*, **F04**, *F05*, **F06.1**, **F06.8**, *G13.8*, **G30.0**, **G30.1**, **G30.8**, **G30.9**, *G31.01, G31.09, G31.1, G31.2, G94, R41.81*, **R54** (any DX on the claim) | *F01.50, F01.51,* **F01.511, F01.518, F01.52, F01.53, F01.54, F01.A0, F01.A11, F01.A18, F01.A2, F01.A3, F01.A4, F01.B0, F01.B11, F01.B18, F01.B2, F01.B3, F01.B4, F01.C0, F01.C11, F01.C18, F01.C2, F01.C3, F01.C4,** *F02.80, F02.81,* **F02.811, F02.818, F02.82, F02.83, F02.84, F02.A0, F02.A11, F02.A18, F02.A2, F02.A3, F02.A4, F02.B0, F02.B11, F02.B18, F02.B2, F02.B3, F02.B4, F02.C0, F02.C11, F02.C18, F02.C2, F02.C3, F02.C4,** *F03.90, F03.91,* **F03.911, F03.918, F03.92, F03.93, F03.94, F03.A0, F03.A11, F03.A18, F03.A2, F03.A3, F03.A4, F03.B0, F03.B11, F03.B18, F03.B2, F03.B3, F03.B4, F03.C0, F03.C11, F03.C18, F03.C2, F03.C3, F03.C4,** *F05, G13.8, G31.01, G31.09, G31.1, G31.2,* ***G31.83,*** *G94, R41.81*(any DX on the claim) | Removed 32 ICD-9 codes; added 70 ICD-10 codes |

| | | |
|---|---|---|
| Reference Period | 3 years | 2 years |
| Number/Type of Claims to Qualify | At least 1 inpatient, SNF, HHA, HOP, or Carrier claim with DX code | At least 1 inpatient/SNF/HHA claim OR 2 HOP/carrier claims with DX codes |

From: https://www2.ccwdata.org/web/guest/condition-categories-chronic

**Notes:* Beginning in 1991, CMS used 27 Chronic Condition Warehouse (CCW) Chronic Condition variables (referred to as "CC27") from validated CCW algorithms. In 2017, CMS began updating and refining these algorithms, resulting in 30 CCW Chronic Condition variables (referred to as "CC30")[1, 2]. By 2022, CC30 had completely replaced CC27. CC27 data is available from 1991-2021, and CC30 data is available from 2017 onward. Between CC27 and CC30, the dementia-related algorithms had undergone some important changes. algorithms for Alzheimer's disease (AD) and Non-Alzheimer's Dementia (referred to as "Alzheimer's Disease, Related Disorders, or Senile Dementia" in CC27 [we continue to refer to this group of conditions as "ADRD"]) saw several important changes: The look-back period for was reduced from 3 to 2 years, and the number of required outpatient or carrier claims increased from 1 to 2[1], and ICD-9 codes were removed entirely. The Alzheimer's Disease and Related Disorders or Senile Dementia (CC27) category was removed entirely and replaced by Non-Alzheimer's Dementia (CC30)[1]. Furthermore, it should be noted that the updated "Non-Alzheimer's Dementia" algorithm does not include any AD-specific ICD codes but does include many non-specific dementia codes (e.g., ICD-9 code 294.8: Other persistent mental disorders) which are frequently applied to individuals with AD-like symptoms [3]. Hence, many individuals with AD are likely classified as "Non- Alzheimer's Dementia" cases according to this algorithm.

A CCW report on the changes between CC27 and CC30 found that the stricter CC30 algorithm reduced the prevalence of non-AD dementia/ADRD by 3.3%[2]. There is also concern that the modifications in look-back period and claims requirement may disproportionately affect identification in certain populations (e.g., racial and ethnic minorities, rural-living individuals)[4], which may obscure important information about ADRD prevalence and diagnoses in these populations.

*In our original paper, we used the CC27 algorithm "Alzheimer's Disease, Related Disorders, or Senile Dementia," which we referred to as "CCW-ADRD." However, the changes between CC27 and CC30 replaced "Alzheimer's Disease, Related Disorders, or Senile Dementia (CC27)" with "Non-Alzheimer's Dementia (CC30)," which is why we included the CCW "Non-Alzheimer's Dementia" code in the current paper.*

**Supplemental Table 2.** List of ICD-9/10 codes for the MVP, PheCode, CCW, and MAP algorithms for AD and ADRD/Dementia used in the original chart review project.

| ICD-9 Codes | ICD-10 Codes |
|---|---|
| 331.0, 294.20, 294.21, 294.8, 290.0, 290.20, 290.21, 290.3, 331.2, 331.7, 290.40, 290.41, 290.42, 290.42, 331.82, 331.1, 331.19, 290.10, 290.11, 290.12, 290.13, 331.5, 333.4, 332, 331.11, 294.10, 294.11, 797 | G30.0, G30.1, G30.8, G30.9, F03.90, F03.91, F01.50, F01.51, G31.83, G31.0, G31.09, G91.2, G10, G20, A81.00, G31.01, G10.96, F02.80, F02.81, G31.1, F04, F05, F06.1, F06.8, G13.8, G31.2, G94, R41,81, R54 |

**Supplemental Table 3.** List of concept unique identifiers (CUIs) used to identify AD-relevant notes to be used in chart review. Any notes identified with a listed CUI based on natural language processing (NLP) were provisioned for chart review.

| Terms | CUI# |
|---|---|
| mental status | C0488568 |
| mental status | C0488569 |
| mental status narrative | C0488568 |
| ad | C1704642 |
| horowitz index | C1977592 |
| acquired immune defic syndrome dementia complex | C0001849 |
| ad | C0002395 |
| ad1 | C1863052 |
| ad10 | C1864828 |
| ad11 | C1853360 |
| ad12 | C1970209 |
| ad13 | C1970147 |
| ad14 | C1970144 |
| ad15 | C1970143 |
| ad16 | C2677888 |
| ad2 | C1863051 |
| ad3 | C1843013 |
| ad4 | C1847200 |
| ad5 | C1865868 |
| ad6 | C1854187 |
| ad7 | C1853555 |
| ad8 | C1846735 |
| ad9 | C1837149 |
| aids dementia | C0001849 |
| aids dementia complex | C0001849 |
| aids relat dementia complex | C0001849 |
| aids related dementia complex | C0001849 |
| alzheimer dementia | C0002395 |
| alzheimer dis | C0002395 |
| alzheimer dis early onset | C0750901 |
| alzheimer disease | C0002395 |
| alzheimer disease 1 | C2931257 |
| alzheimer disease 10 | C1864828 |
| alzheimer disease 11 | C1853360 |
| alzheimer disease 12 | C1970209 |
| alzheimer disease 13 | C1970147 |
| alzheimer disease 14 | C1970144 |
| alzheimer disease 15 | C1970143 |
| alzheimer disease 16 | C2677888 |
| alzheimer disease 2 | C1863051 |
| alzheimer disease 3 | C1843013 |
| alzheimer disease 4 | C1847200 |
| alzheimer disease 5 | C1865868 |
| alzheimer disease 6 | C1854187 |
| alzheimer disease 7 | C1853555 |
| alzheimer disease 8 | C1846735 |
| alzheimer disease 9 | C1837149 |
| alzheimer disease associated with apoe e4 | C1863051 |
| alzheimer disease associated with apoe4 | C1863051 |
| alzheimer disease early onset | C0750901 |
| alzheimer disease early onset type 3 | C1843013 |
| alzheimer disease type 1 | C2931257 |
| alzheimer disease type 2 | C1863051 |
| alzheimer disease type 3 | C1843013 |
| alzheimer disease type 4 | C1847200 |
| alzheimer disease without neurofibrillary tangles | C1858751 |
| alzheimer diseases | C0002395 |
| alzheimer sclerosis | C0002395 |
| alzheimer syndrome | C0002395 |
| alzheimer type dementia | C0002395 |
| alzheimer type senile dementia | C0002395 |
| alzheimer's dementia | C0002395 |
| alzheimer's disease | C0002395 |
| alzheimer's disease early onset | C0750901 |
| alzheimer's disease without neurofibrillary tangles | C1858751 |
| alzheimer's disease without tau pathology | C1858751 |
| alzheimer's diseases | C0002395 |
| alzheimers dementia | C0002395 |

| | |
|---|---|
| alzheimers dis | C0002395 |
| alzheimers disease | C0002395 |
| alzheimers disease early onset | C0750901 |
| arteriosclerotic dementia | C0600359 |
| arteriosclerotic dementia nos | C0600359 |
| arteriosclerotic dementias | C0600359 |
| ataxia with myoclonic epilepsy and presenile dementia | C1859646 |
| atherosclerotic dementia | C0600359 |
| bodies dementia lewy | C0752347 |
| bodies dementias lewy | C0752347 |
| bodies disease lewy | C0752347 |
| body dementia lewi | C0752347 |
| body dementia lewis | C0752347 |
| body dementia lewy | C0752347 |
| body disease lewi | C0752347 |
| body disease lewis | C0752347 |
| body disease lewy | C0752347 |
| body diseases lewy | C0752347 |
| body lewy dementia | C0752347 |
| catatonia of kraepelin | C1867772 |
| dat | C0002395 |
| dats | C0002395 |
| ddpac | C0338451 |
| dementia aids | C0001849 |
| dementia alzheimers | C0002395 |
| dementia complex acquired immune defic syndrome | C0001849 |
| dementia complex aids relat | C0001849 |
| dementia frontal | C1260405 |
| dementia frontotemporal | C0338451 |
| dementia hiv | C0001849 |
| dementia infarct multi | C0011263 |
| dementia lewy bodies | C0752347 |
| dementia multi infarct | C0011263 |
| dementia multiinfarct | C0011263 |
| dementia of the alzheimer type | C0002395 |
| dementia of the alzheimer's type | C0002395 |

| | |
|---|---|
| dementia senile/alzheimer | C0877811 |
| dementia vascular | C0011269 |
| dementia w lewy bodies | C0752347 |
| dementia with lewy bodies | C0752347 |
| diffuse lewy body dis | C0752347 |
| diffuse lewy body disease | C0752347 |
| disinhibition dementia parkinsonism amyotrophy complex | C0338451 |
| disinhibition dementia parkinsonism amytrophy complex | C0338451 |
| dlb | C0752347 |
| early onset alzheimer dis | C0750901 |
| early onset alzheimer disease | C0750901 |
| early onset alzheimer's disease | C0750901 |
| familial alzheimer disease | C0276496 |
| familial alzheimer's disease | C0276496 |
| familial danish dementia | C1861735 |
| familial pick disease | C0338451 |
| familial pick's disease | C0338451 |
| familial pick's diseases | C0338451 |
| familial picks disease | C0338451 |
| fdd | C1861735 |
| fldem | C0338451 |
| frontal dementia | C1260405 |
| frontal lobe dementia | C0338455 |
| frontotemporal dementia | C0338451 |
| frontotemporal dementia with parkinsonism | C0338451 |
| frontotemporal dementia with parkinsonism 17 | C0338451 |
| frontotemporal dementias | C0338451 |
| frontotemporal lobar degeneration with tau inclusions | C0338451 |
| frontotemporal lobar degeneration with ubiquitin positive inclusions | C0338451 |
| frontotemporal lobe dementia | C0338451 |

| | |
|---|---|
| frontotemporal lobe dementias | C0338451 |
| ftd | C0338451 |
| ftdp17 | C0338451 |
| ftld with tau inclusions | C0338451 |
| ftld with tdp 43 pathology | C0338451 |
| grn related frontotemporal dementia | C0338451 |
| hd | C0020179 |
| hddd1 | C0338451 |
| hddd2 | C0338451 |
| hdl2 | C1847987 |
| hereditary dysphasic disinhibition dementia | C0338451 |
| heredopathia ophthalmootoencephalica | C1861735 |
| hiv 01 associated cognitive motor complex | C0936243 |
| hiv 1 assoc cognitive motor complex | C0936243 |
| hiv 1 associated cognitive motor complex | C0936243 |
| hiv 1 cognitive and motor complex | C0936243 |
| hiv assoc cognitive motor complex | C0001849 |
| hiv associated cognitive and motor complex | C0001849 |
| hiv associated cognitive motor complex | C0001849 |
| hiv dementia | C0001849 |
| hiv dementias | C0001849 |
| hooe | C1861735 |
| huntington chorea | C0020179 |
| huntington chronic progressive hereditary chorea | C0020179 |
| huntington dis | C0020179 |
| huntington disease | C0020179 |
| huntington's chorea | C0020179 |
| huntington's disease | C0020179 |
| huntingtons chorea | C0020179 |
| huntingtons dis | C0020179 |
| huntingtons disease | C0020179 |
| late onset familial alzheimer disease | C1863051 |

| | |
|---|---|
| lewy bodies dementia | C0752347 |
| lewy body dementia | C0752347 |
| lewy body dis | C0752347 |
| lewy body dis cortical | C0752347 |
| lewy body dis diffuse | C0752347 |
| lewy body disease | C0752347 |
| lewy body type senile dementia | C0752347 |
| lewy body variant of alzheimer disease | C1851958 |
| mast syndrome | C1855346 |
| motor neuron disease with dementia and ophthalmoplegia | C1838253 |
| mstd | C0338451 |
| multi infarct dementia | C0011263 |
| multifocal dementia | C0011263 |
| multiinfarct dementia | C0011263 |
| multiinfarct dementias | C0011263 |
| multiple system tauopathy with presenile dementia | C0338451 |
| opticoacoustic nerve atrophy with dementia | C1839564 |
| opticoacustic nerve atrophy with dementia | C1839564 |
| presenile alzheimer dementia | C0750901 |
| primary senile degenerative dementia | C0002395 |
| sdat | C0002395 |
| senile dementia | C0002395 |
| senile dementia of the alzheimer type | C0002395 |
| sensory neuropathy with deafness and dementia | C2931309 |
| simple senile dementia | C0002395 |
| spg21 | C1855346 |
| subcortical dementia | C4024935 |
| syndrome of opticoacoustic nerve atrophy with dementia | C1839564 |
| vascular dementia | C0011269 |
| vascular dementias | C0011269 |
| wilhelmsen lynch disease | C0338451 |
| wld | C0338451 |
| wright dyck syndrome | C2931309 |

| | |
|---|---|
| cerebral atrophy diffuse | C0598275 |
| clinical findings relating to memory | C0700327 |
| ctcae grade 2 mental status | C1561354 |
| ctcae grade 3 mental status | C1561355 |
| diffuse cerebral atrophy | C0598275 |
| frontolimbic dementia | C1836151 |
| grade 2 mental status | C1561354 |
| grade 3 mental status | C1561355 |
| memories | C0700327 |
| memory | C0700327 |
| memory observations | C0700327 |
| mental state | C0278060 |
| mental status | C0278060 |
| mental status | C2004284 |
| mental status adverse event | C2004284 |
| motor deterioration | C1866284 |
| neural tube defect risk | C1954450 |
| neurologic deterioration | C1854838 |
| progressive mental deterioration | C1854838 |
| progressive neurodegeneration | C1854838 |
| progressive neurologic deterioration | C1854838 |
| psychomotor degeneration | C1836842 |
| psychomotor deterioration | C1836842 |
| social and occupational deterioration | C1866986 |
| cognitive | C1516691 |
| dictated | C1548384 |
| verbal | C1548941 |
| ad | C0002838 |
| verbal | C1608381 |
| cue | C0010439 |
| cueing | C0010439 |
| cues | C0010439 |
| ad | C1578437 |
| ad | C1706476 |
| ad term type | C1706476 |
| memory | C1706377 |
| persistent memory | C1882350 |

| | |
|---|---|
| persistent memory device | C1882350 |
| acute confusional senile dementia | C0546126 |
| alcohol or other drug related dementia | C0679298 |
| alzheimer dis late onset | C0494463 |
| alzheimers dis focal onset | C0750900 |
| amentia | C0497327 |
| amentias | C0497327 |
| aodr dementia | C0679298 |
| bad memory | C0233794 |
| cognitive decline | C0338656 |
| cognitive defects | C0338656 |
| cognitive deficits | C0338656 |
| cognitive disturbance | C0338656 |
| cognitive dysfunction | C0338656 |
| cognitive dysfunctions | C0338656 |
| cognitive impairment | C0338656 |
| cognitive impairments | C0338656 |
| demen nos w behav distrb | C3161079 |
| demen nos w/o behv dstrb | C3161078 |
| dementia | C0011265 |
| dementia | C0497327 |
| dementia acquired | C0234984 |
| dementia disorder | C0497327 |
| dementia disorders | C0497327 |
| dementia due to specified medical condition | C0236752 |
| dementia in conditions classified elsewhere | C0338632 |
| dementia in conditions classified elsewhere nos | C0338632 |
| dementia nos | C0497327 |
| dementia presenile | C0011265 |
| dementia progressive | C0497327 |
| dementia secondary | C0856416 |
| dementia senile | C0011268 |
| dementias | C0497327 |
| diffuse neurofibrillary tangles with calcification | C2717758 |
| disturb memory/concentration | C0233794 |
| disturbance memory/concentration | C0233794 |
| familial dementia | C0751071 |

| | |
|---|---|
| familial dementias | C0751071 |
| focal onset alzheimer's disease | C0750900 |
| focal onset alzheimers dis | C0750900 |
| frontotemporal lobar degeneration | C0751072 |
| frontotemporal lobar degenerations | C0751072 |
| ftld | C0751072 |
| ftlds | C0751072 |
| impair memory | C0233794 |
| impaired cognition | C0338656 |
| impaired memory | C0233794 |
| impairment memory | C0233794 |
| impairments memory | C0233794 |
| kluver bucy syndrome | C0270707 |
| kosaka shibayama disease | C2717758 |
| late onset alzheimer dis | C0494463 |
| late onset alzheimer disease | C0494463 |
| mci | C1270972 |
| memory deficit | C0233794 |
| memory deficits | C0233794 |
| memory disturbance | C0233794 |
| memory impaired | C0233794 |
| memory impairment | C0233794 |
| memory poor | C0233794 |
| memory problem | C0233794 |
| memory problems | C0233794 |
| mesulam syndrome | C0282513 |
| mesulam's syndrome | C0282513 |
| mild cognitive disorder | C1270972 |
| mild cognitive impairment | C1270972 |
| mild cognitive impairments | C1270972 |
| neurocognitive disturbance | C0338656 |
| neurocognitive dysfunction | C0338656 |
| other specified senile psychotic conditions | C0154319 |
| poor memory | C0233794 |
| ppa | C0282513 |
| pre senile dementia | C0011265 |

| | |
|---|---|
| presenile and senile dementia | C1541844 |
| presenile delirium | C0154309 |
| presenile delusion | C0154310 |
| presenile dementia | C0011265 |
| presenile dementia | C0677545 |
| presenile dementia nos | C0011265 |
| presenile dementia with delirium | C0154309 |
| presenile dementia with delusional features | C0154310 |
| presenile dementia with depressive features | C0338629 |
| presenile depression | C0338629 |
| presenile organic psychotic conditions | C0011265 |
| primary progressive aphasia | C0282513 |
| primary progressive aphasias | C0282513 |
| progressive dementia | C0497327 |
| progressive primary aphasia | C0282513 |
| secondary dementia | C0856416 |
| senile delirium | C0154315 |
| senile dementia | C0011268 |
| senile dementia nos | C0011268 |
| senile dementia uncomp | C3665587 |
| senile dementia with acute confusional state | C0154315 |
| senile dementia with delirium | C0154315 |
| senile dementia with delusional or depressive features | C0859643 |
| senile dementia with depressive features | C0338631 |
| senile depressive | C0338631 |
| senile paranoid dementia | C0338630 |
| senile paranoid dementias | C0338630 |
| senile psychosis nec | C0154319 |
| senile psychosis nos | C1457889 |
| senile psychot cond nos | C1457889 |
| unspecified senile psychotic condition | C1457889 |
| autobiographical memories | C0561843 |

| | |
|---|---|
| autobiographical memory | C0561843 |
| behavioral habituation/sensitization | C0178500 |
| characteristic of memory | C0679059 |
| cognition | C0009240 |
| cognition/thought processes | C0009240 |
| cognitions | C0009240 |
| cognitive function | C0009240 |
| cognitive functions | C0009240 |
| comprehension | C0162340 |
| episodic memories | C0561843 |
| episodic memory | C0561843 |
| experience | C0596545 |
| experiences | C0596545 |
| immediate memories | C0025265 |
| immediate memory | C0025265 |
| long term memory | C0423909 |
| longterm memories | C0423909 |
| longterm memory | C0423909 |
| memory | C0025260 |
| memory by content | C0679050 |
| memory function | C0025260 |
| memory interference | C0679066 |
| memory photogr | C0013727 |
| memory process | C0679052 |
| memory processing | C0679052 |
| memory recognition | C0524637 |
| memory short term | C0025265 |
| memory spatial | C0814087 |
| memory work | C0025265 |
| memory working | C0025265 |
| mental | C0229992 |
| mental association | C0004083 |
| mental process | C0229992 |
| mental recall | C0034770 |
| mtm | C3156335 |

| | |
|---|---|
| photogr memory | C0013727 |
| photographic memories | C0013727 |
| photographic memory | C0013727 |
| process memory | C0679052 |
| recall | C0034770 |
| recalling | C0034770 |
| recognition | C0524637 |
| recognition memory | C0524637 |
| recollect | C0034770 |
| recollection | C0034770 |
| remember | C0034770 |
| remembering | C0034770 |
| remembers | C0034770 |
| repetition priming | C3179029 |
| response generalization | C0017325 |
| response generalizations | C0017325 |
| retention | C0035280 |
| retention psychol | C0035280 |
| short term memory | C0025265 |
| shortterm memories | C0025265 |
| shortterm memory | C0025265 |
| spatial memories | C0814087 |
| spatial memory | C0814087 |
| verbal memory | C0561770 |
| visual learning | C0582587 |
| visual memory | C0542316 |
| working memories | C0025265 |
| working memory | C0025265 |
| kengshenmycin | C0647391 |
| cognitive decline | C0234985 |
| intellectual deterioration | C0234985 |
| mental deterioration | C0234985 |
| progressive cognitive decline | C0234985 |
| ad | C0001555 |
| ad | C0547043 |

**Supplemental Table 4.** Algorithm performance in MVP participants of European ancestry in terms of association with the *APOE* ε4 locus.

| **Algorithm** | ***APOE* ε4 OR (CI)** | **Z- value** |
|---|---|---|
| MVP-AD (VA EMR) | 2.40 (2.31,2.50) | 44.85539 |
| MVP-AD (VA EMR) 60+ | 2.50 (2.40,2.60) | 46.0388 |
| MVP-AD (VA EMR) 65+ | 2.53 (2.43,2.64) | 45.41892 |
| MVP-AD (VA EMR/CMS) | 2.08 (2.02,2.14) | 51.49583 |
| MVP-AD (VA EMR/CMS) 60+ | 2.17 (2.11,2.23) | 53.18244 |
| MVP-AD (VA EMR/CMS) 65+ | 2.20 (2.14,2.27) | 52.83065 |
| CCW-AD (VA EMR) | 2.33 (2.25,2.42) | 46.25662 |
| CCW-AD (VA EMR) 60+ | 2.42 (2.33,2.51) | 47.42518 |
| CCW-AD (VA EMR) 65+ | 2.47 (2.38,2.56) | 47.46174 |
| CCW-AD (VA EMR/CMS) | 2.03 (1.97,2.08) | 52.03586 |
| CCW-AD (VA EMR/CMS) 60+ | 2.10 (2.04,2.16) | 53.70773 |
| CCW-AD (VA EMR/CMS) 65+ | 2.15 (2.09,2.21) | 54.12314 |
| Phe-AD (VA EMR) | 2.41 (2.32,2.50) | 45.42059 |
| Phe-AD (VA EMR) 60+ | 2.50 (2.40,2.60) | 46.58311 |
| Phe-AD (VA EMR) 65+ | 2.54 (2.45,2.65) | 46.5169 |
| Phe-AD (VA EMR/CMS) | 2.08 (2.02,2.13) | 51.78383 |
| Phe-AD (VA EMR/CMS) 60+ | 2.15 (2.09,2.22) | 53.43818 |
| Phe-AD (VA EMR/CMS) 65+ | 2.20 (2.13,2.26) | 53.78054 |
| MVP-AD+ (VA EMR) | 1.83 (1.79,1.87) | 53.74432 |
| MVP-AD+ (VA EMR)60+ | 1.92 (1.88,1.97) | 55.86316 |
| MVP-AD+ (VA EMR) 65+ | 1.97 (1.93,2.02) | 55.67395 |
| MVP-AD+ (VA EMR/CMS) | 1.64 (1.61,1.67) | 53.41544 |
| MVP-AD+ (VA EMR/CMS) 60+ | 1.73 (1.69,1.76) | 55.88085 |
| MVP-AD+ (VA EMR/CMS) 65+ | 1.77 (1.73,1.80) | 55.93393 |
| MVP-ADRD (VA EMR) | 1.77 (1.73,1.81) | 54.04005 |
| MVP-ADRD (VA EMR) 60+ | 1.86 (1.82,1.91) | 56.44191 |
| MVP-ADRD (VA EMR) 65+ | 1.92 (1.87,1.96) | 56.49455 |
| MVP-ADRD (VA EMR/CMS) | 1.60 (1.58,1.63) | 52.55449 |
| MVP-ADRD (VA EMR/CMS) 60+ | 1.68 (1.65,1.71) | 55.19444 |
| MVP-ADRD (VA EMR/CMS) 65+ | 1.72 (1.69,1.76) | 55.42377 |
| MVP-Dementia (VA EMR) | 1.54 (1.52.1.57) | 45.92709 |
| MVP-Dementia (VA EMR) 60+ | 1.63 (1.59,1.66) | 48.68267 |
| MVP-Dementia (VA EMR) 65+ | 1.68 (1.64,1.71) | 49.36537 |
| MVP-Dementia (VA EMR/CMS) | 1.47 (1.44,1.49) | 45.66526 |
| MVP-Dementia (VA EMR/CMS) 60+ | 1.54 (1.51,1.56) | 48.7174 |
| MVP-Dementia (VA EMR/CMS) 65+ | 1.58 (1.55,1.61) | 49.45364 |
| CCW-Non-AD Dementia (VA EMR) | 1.71 (1.68,1.74) | 51.88177 |
| CCW-Non-AD Dementia (VA EMR) 60+ | 1.77 (1.74,1.81) | 53.80979 |
| CCW-Non-AD Dementia (VA EMR) 65+ | 1.81 (1.77,1.85) | 54.29192 |

| | | |
|---|---|---|
| CCW-Non-AD Dementia (VA EMR/CMS) | 1.54 (1.51,1.56) | 49.9686 |
| CCW-Non-AD Dementia (VA EMR/CMS) 60+ | 1.59 (1.57,1.62) | 52.28936 |
| CCW-Non-AD Dementia (VA EMR/CMS) 65+ | 1.62 (1.60,1.66) | 52.9923 |
| Phe-Dementia (VA EMR) | 1.79 (1.75,1.83) | 54.79118 |
| Phe-Dementia (VA EMR) 60+ | 1.87 (1.83,1.91) | 57.1601 |
| Phe-Dementia (VA EMR) 65+ | 1.91 (1.87,1.96) | 57.25418 |
| Phe-Dementia (VA EMR/CMS) | 1.60 (1.58,1.63) | 52.68302 |
| Phe-Dementia (VA EMR/CMS) 60+ | 1.68 (1.64,1.71) | 55.37621 |
| Phe-Dementia (VA EMR/CMS) 65+ | 1.71 (1.68,1.74) | 55.70434 |
| AD Meds (VA EMR/CMS) | 1.84 (1.80,1.88) | 52.5095 |
| AD Meds (VA EMR/CMS) 60+ | 1.96 (1.91,2.01) | 55.39478 |
| AD Meds (VA EMR/CMS) 65+ | 2.02 (1.97,2.07) | 55.55343 |
| AD Meds/ CCW Non-AD Dementia (VA EMR/ CMS) | 1.52 (1.50,1.54) | 50.95725 |
| AD Meds/CCW Non-AD Dementia (VA EMR/ CMS) 60+ | 1.58 (1.56,1.61) | 53.58836 |
| AD Meds/ CCW Non-AD Dementia (VA EMR/ CMS) 65+ | 1.62 (1.59,1.65) | 54.5493 |
| AD Meds/ MVP-Dementia (VA EMR/ CMS) | 1.46 (1.44,1.49) | 46.8367 |
| AD Meds/ MVP-Dementia (VA EMR/ CMS) 60+ | 1.54 (1.51,1.56) | 50.12438 |
| AD Meds/ MVP-Dementia (VA EMR/ CMS) 65+ | 1.58 (1.55,1.61) | 51.06884 |
| AD Meds/MVP-ADRD (VA EMR/CMS) | 1.58 (1.55,1.60) | 53.04986 |
| AD Meds/MVP-ADRD (VA EMR/CMS) 60+ | 1.65 (1.63,1.69) | 56.05057 |
| AD Meds/MVP-ADRD (VA EMR/CMS) 65+ | 1.70 (1.67,1.73) | 56.60407 |
| AD Meds/ Phe-Dementia (VA EMR/CMS) | 1.57 (1.54,1.60) | 52.85931 |
| AD Meds/ Phe-Dementia (VA EMR/CMS) 60+ | 1.65 (1.62,1.68) | 55.81077 |
| AD Meds/ Phe-Dementia (VA EMR/CMS) 65+ | 1.69 (1.66,1.71) | 56.50235 |
| AD Meds/ MVP-AD+ (VA EMR/CMS) | 1.61 (1.58,1.63) | 53.95184 |
| AD Meds/ MVP-AD+ (VA EMR/CMS) 60+ | 1.69 (1.66,1.72) | 56.8701 |
| AD Meds/ MVP-AD+ (VA EMR/CMS) 65+ | 1.73 (1.70,1.77) | 57.2982 |
| AD Meds/ CCW-AD (VA EMR/ CMS) | 1.79 (1.76,1.83) | 56.40511 |
| AD Meds/ CCW-AD (VA EMR/ CMS) 60+ | 1.89 (1.86,1.94) | 59.20662 |
| AD Meds/ CCW-AD (VA EMR/ CMS) 65+ | 1.95 (1.91,2.00) | 59.86545 |
| AD Meds/ Phe-AD (VA EMR/CMS) | 1.81 (1.77,1.84) | 56.39624 |
| AD Meds/ Phe-AD (VA EMR/CMS) 60+ | 1.91 (1.87,1.95) | 59.25553 |
| AD Meds/ Phe-AD (VA EMR/CMS) 65+ | 1.97 (1.93,2.01) | 59.8747 |
| AD Meds/ MVP-AD (VA EMR/CMS) | 1.81 (1.77,1.85) | 56.33665 |
| AD Meds/ MVP-AD (VA EMR/CMS) 60+ | 1.91 (1.87,1.96) | 59.21984 |
| AD Meds/ MVP-AD (VA EMR/CMS) 65+ | 1.97 (1.93,2.01) | 59.45195 |
| AD Meds (VA EMR) | 1.83 (1.79,1.87) | 52.11468 |
| AD Meds (VA EMR) 60+ | 1.94 (1.89,1.98) | 54.69133 |
| AD Meds (VA EMR) 65+ | 1.99 (1.95,2.04) | 54.77259 |
| AD Meds/ CCW Non-AD Dementia (VA EMR) | 1.65 (1.62,1.68) | 53.83933 |
| AD Meds/CCW Non-AD -Dementia (VA EMR) 60+ | 1.73 (1.69,1.76) | 56.19742 |
| AD Meds/ CCW Non-AD Dementia (VA EMR) 65+ | 1.77 (1.74,1.81) | 57.02803 |
| AD Meds/ MVP-Dementia (VA EMR) | 1.53 (1.50,1.56) | 47.58335 |

| | | |
|---|---|---|
| AD Meds/ MVP-Dementia (VA EMR) 60+ | 1.61 (1.58,1.64) | 50.56031 |
| AD Meds/ MVP-Dementia (VA EMR) 65+ | 1.66 (1.63,1.69) | 51.48747 |
| AD Meds/ MVP-ADRD (VA EMR) | 1.69 (1.66,1.72) | 54.49345 |
| AD Meds/ MVP-ADRD (VA EMR) 60+ | 1.78 (1.75,1.82) | 57.30115 |
| AD Meds/ MVP-ADRD (VA EMR) 65+ | 1.83 (1.80,1.87) | 57.75222 |
| AD Meds/ Phe-Dementia (VA EMR) | 1.69 (1.66,1.73) | 54.75439 |
| AD Meds/ Phe-Dementia (VA EMR) 60+ | 1.78 (1.75,1.82) | 57.51427 |
| AD Meds/ Phe-Dementia (VA EMR) 65+ | 1.83 (1.80,1.87) | 58.09743 |
| AD Meds/ MVP- AD+ (VA EMR) | 1.71 (1.68,1.75) | 54.60913 |
| AD Meds/ MVP- AD+ (VA EMR) 60+ | 1.81 (1.77,1.85) | 57.29041 |
| AD Meds/ MVP- AD+ (VA EMR) 65+ | 1.86 (1.82,1.90) | 57.60037 |
| AD Meds/ CCW-AD (VA EMR) | 1.82 (1.78,1.86) | 53.71742 |
| AD Meds/ CCW-AD (VA EMR) 60+ | 1.92 (1.88,1.97) | 56.26835 |
| AD Meds/ CCW-AD (VA EMR) 65+ | 1.98 (1.94,2.03) | 56.68243 |
| AD Meds/ Phe-AD (VA EMR) | 1.83 (1.79,1.87) | 53.48161 |
| AD Meds/ Phe-AD (VA EMR) 60+ | 1.93 (1.89,1.97) | 56.06918 |
| AD Meds/ Phe-AD (VA EMR) 65+ | 1.99 (1.94,2.04) | 56.46082 |
| AD Meds/ MVP-AD (VA EMR) | 1.83 (1.79,1.87) | 53.40863 |
| AD Meds/ MVP-AD (VA EMR) 60+ | 1.93 (1.89,1.97) | 55.97115 |
| AD Meds/ MVP-AD (VA EMR) 65+ | 1.98 (1.94,2.03) | 56.11379 |

**Supplemental Table 5.** Algorithm performance in MVP participants of European ancestry in terms of association with an AD polygenic risk score (PRS) based on the Kunkle et al. (2019) AD GWAS excluding the chromosome 19 *APOE* region. OR is in terms of risk per 1 standard deviation increase in the PRS.

| **Algorithms** | **PRS OR (CI)** | **Z-value** | **P-Value** |
|---|---|---|---|
| MVP-AD (VA EMR) | 1.19 (1.16,1.22) | 14.64 | 1.46E-48 |
| MVP-AD (VA EMR) 60+ | 1.20 (1.17,1.23) | 15.07 | 2.37E-51 |
| MVP-AD (VA EMR) 65+ | 1.20 (1.17,1.23) | 14.70 | 6.45E-49 |
| MVP-AD (VA EMR/CMS) | 1.15 (1.13,1.17) | 16.98 | 1.20E-64 |
| MVP-AD (VA EMR/CMS) 60+ | 1.16 (1.14,1.18) | 17.56 | 5.01E-69 |
| MVP-AD (VA EMR/CMS) 65+ | 1.16 (1.14,1.18) | 17.27 | 7.72E-67 |
| CCW-AD (VA EMR) | 1.18 (1.16,1.21) | 14.85 | 6.75E-50 |
| CCW-AD (VA EMR) 60+ | 1.19 (1.16,1.22) | 15.36 | 3.02E-53 |
| CCW-AD (VA EMR) 65+ | 1.19 (1.17,1.22) | 15.29 | 9.47E-53 |
| CCW-AD (VA EMR/CMS) | 1.14 (1.12,1.16) | 16.71 | 1.03E-62 |
| CCW-AD (VA EMR/CMS) 60+ | 1.15 (1.13,1.17) | 17.45 | 3.72E-68 |
| CCW-AD (VA EMR/CMS) 65+ | 1.15 (1.13,1.17) | 17.34 | 2.48E-67 |
| Phe-AD (VA EMR) | 1.19 (1.16,1.22) | 14.54 | 6.35E-48 |
| Phe-AD (VA EMR) 60+ | 1.20 (1.17,1.22) | 14.94 | 1.89E-50 |
| Phe-AD (VA EMR) 65+ | 1.20 (1.17,1.23) | 14.89 | 4.09E-50 |
| Phe-AD (VA EMR/CMS) | 1.15 (1.13,1.17) | 17.05 | 3.55E-65 |
| Phe-AD (VA EMR/CMS) 60+ | 1.16 (1.14,1.18) | 17.72 | 3.21E-70 |
| Phe-AD (VA EMR/CMS) 65+ | 1.16 (1.14,1.18) | 17.62 | 1.64E-69 |
| MVP-AD+ (VA EMR) | 1.11 (1.10,1.13) | 16.83 | 1.57E-63 |
| MVP-AD+ (VA EMR)60+ | 1.12 (1.11,1.14) | 17.56 | 5.33E-69 |
| MVP-AD+ (VA EMR) 65+ | 1.13 (1.11,1.15) | 17.63 | 1.55E-69 |
| MVP-AD+ (VA EMR/CMS) | 1.09 (1.08,1.10) | 16.81 | 2.02E-63 |
| MVP-AD+ (VA EMR/CMS) 60+ | 1.10 (1.08,1.11) | 17.56 | 5.10E-69 |
| MVP-AD+ (VA EMR/CMS) 65+ | 1.10 (1.09,1.11) | 17.75 | 1.77E-70 |
| MVP-ADRD (VA EMR) | 1.11 (1.09,1.12) | 16.86 | 8.86E-64 |
| MVP-ADRD (VA EMR) 60+ | 1.11 (1.10,1.13) | 17.43 | 4.72E-68 |
| MVP-ADRD (VA EMR) 65+ | 1.12 (1.11,1.14) | 17.62 | 1.71E-69 |
| MVP-ADRD (VA EMR/CMS) | 1.08 (1.07,1.09) | 16.56 | 1.33E-61 |
| MVP-ADRD (VA EMR/CMS) 60+ | 1.09 (1.08,1.10) | 17.23 | 1.69E-66 |
| MVP-ADRD (VA EMR/CMS) 65+ | 1.10 (1.09,1.11) | 17.46 | 2.91E-68 |
| MVP-Dementia (VA EMR) | 1.08 (1.07,1.09) | 15.05 | 3.05E-51 |
| MVP-Dementia (VA EMR) 60+ | 1.09 (1.08,1.10) | 15.90 | 6.10E-57 |
| MVP-Dementia (VA EMR) 65+ | 1.10 (1.08,1.11) | 16.10 | 2.37E-58 |
| MVP-Dementia (VA EMR/CMS) | 1.07 (1.06,1.08) | 15.35 | 3.39E-53 |
| MVP-Dementia (VA EMR/CMS) 60+ | 1.08 (1.07,1.09) | 16.30 | 1.03E-59 |
| MVP-Dementia (VA EMR/CMS) 65+ | 1.08 (1.07,1.09) | 16.48 | 5.49E-61 |
| CCW-Non-AD Dementia (VA EMR) | 1.10 (1.08,1.11) | 15.96 | 2.20E-57 |
| CCW-Non-AD Dementia (VA EMR) 60+ | 1.10 (1.09,1.12) | 16.77 | 3.56E-63 |

| | | | |
|---|---|---|---|
| CCW-Non-AD Dementia (VA EMR) 65+ | 1.11 (1.09,1.12) | 16.85 | 1.09E-63 |
| CCW-Non-AD Dementia (VA EMR/CMS) | 1.07 (1.06,1.08) | 14.83 | 9.71E-50 |
| CCW-Non-AD Dementia (VA EMR/CMS) 60+ | 1.08 (1.07,1.09) | 15.76 | 5.50E-56 |
| CCW-Non-AD Dementia (VA EMR/CMS) 65+ | 1.08 (1.07,1.09) | 15.90 | 5.89E-57 |
| Phe-Dementia (VA EMR) | 1.11 (1.10,1.12) | 17.28 | 6.94E-67 |
| Phe-Dementia (VA EMR) 60+ | 1.12 (1.10,1.13) | 17.91 | 9.32E-72 |
| Phe-Dementia (VA EMR) 65+ | 1.12 (1.11,1.14) | 18.01 | 1.55E-72 |
| Phe-Dementia (VA EMR/CMS) | 1.08 (1.07,1.09) | 16.53 | 2.24E-61 |
| Phe-Dementia (VA EMR/CMS) 60+ | 1.09 (1.08,1.10) | 17.32 | 3.27E-67 |
| Phe-Dementia (VA EMR/CMS) 65+ | 1.09 (1.08,1.11) | 17.48 | 2.15E-68 |
| AD Meds (VA EMR/CMS) | 1.11 (1.09,1.13) | 16.13 | 1.60E-58 |
| AD Meds (VA EMR/CMS) 60+ | 1.13 (1.11,1.14) | 17.45 | 3.55E-68 |
| AD Meds (VA EMR/CMS) 65+ | 1.14 (1.12,1.15) | 17.68 | 6.51E-70 |
| AD Meds/ CCW Non-AD Dementia (VA EMR/ CMS) | 1.07 (1.06,1.08) | 14.87 | 5.30E-50 |
| AD Meds/CCW Non-AD Dementia (VA EMR/ CMS) 60+ | 1.08 (1.07,1.09) | 16.14 | 1.42E-58 |
| AD Meds/ CCW Non-AD Dementia (VA EMR/ CMS) 65+ | 1.08 (1.07,1.09) | 16.38 | 2.77E-60 |
| AD Meds/ MVP-Dementia (VA EMR/CMS) | 1.07 (1.06, 1.08) | 15.18 | 4.99E-52 |
| AD Meds/ MVP-Dementia (VA EMR/CMS) 60+ | 1.08 (1.07,1.09) | 16.59 | 7.77E-62 |
| AD Meds/ MVP-Dementia (VA EMR/CMS) 65+ | 1.08 (1.07,1.09) | 16.88 | 6.69E-64 |
| AD Meds/MVP-ADRD (VA EMR/CMS) | 1.08 (1.07,1.09) | 16.40 | 1.90E-60 |
| AD Meds/MVP-ADRD (VA EMR/CMS) 60+ | 1.09 (1.08,1.10) | 17.55 | 5.89E-69 |
| AD Meds/MVP-ADRD (VA EMR/CMS) 65+ | 1.09 (1.08,1.10) | 17.89 | 1.29E-71 |
| AD Meds/ Phe-Dementia (VA EMR/CMS) | 1.08 (1.07,1.09) | 16.28 | 1.40E-59 |
| AD Meds/ Phe-Dementia (VA EMR/CMS) 60+ | 1.09 (1.08,1.10) | 17.42 | 5.49E-68 |
| AD Meds/ Phe-Dementia (VA EMR/CMS) 65+ | 1.09 (1.08,1.10) | 17.64 | 1.11E-69 |
| AD Meds/ MVP- AD+ (VA EMR/ CMS) | 1.08 (1.07,1.09) | 17.10 | 1.60E-65 |
| AD Meds/ MVP- AD+ (VA EMR/ CMS) 60+ | 1.09 (1.08,1.10) | 18.02 | 1.46E-72 |
| AD Meds/ MVP- AD+ (VA EMR/ CMS) 65+ | 1.10 (1.09,1.11) | 18.41 | 1.19E-75 |
| AD Meds/ CCW-AD (VA EMR/ CMS) | 1.11 (1.10,1.12) | 16.72 | 9.45E-63 |
| AD Meds/ CCW-AD (VA EMR/ CMS) 60+ | 1.12 (1.11,1.14) | 19.43 | 4.38E-84 |
| AD Meds/ CCW-AD (VA EMR/ CMS) 65+ | 1.13 (1.12,1.14) | 19.63 | 8.92E-86 |
| AD Meds/ Phe-AD (VA EMR/CMS) | 1.11 (1.10,1.13) | 18.02 | 1.46E-72 |
| AD Meds/ Phe-AD (VA EMR/CMS) 60+ | 1.13 (1.11,1.14) | 19.41 | 6.27E-84 |
| AD Meds/ Phe-AD (VA EMR/CMS) 65+ | 1.13 (1.12,1.15) | 19.53 | 5.80E-85 |
| AD Meds/ MVP-AD (VA EMR/CMS) | 1.11 (1.10,1.12) | 18.01 | 1.63E-72 |
| AD Meds/ MVP-AD (VA EMR/CMS) 60+ | 1.13 (1.11,1.14) | 19.36 | 1.54E-83 |
| AD Meds/ MVP-AD (VA EMR/CMS) 65+ | 1.13 (1.12,1.15) | 19.53 | 5.80E-85 |
| AD Meds (VA EMR) | 1.11 (1.10,1.13) | 16.06 | 4.65E-58 |
| AD Meds (VA EMR) 60+ | 1.13 (1.11,1.14) | 17.33 | 2.93E-67 |
| AD Meds (VA EMR) 65+ | 1.13 (1.12,1.15) | 17.56 | 4.91E-69 |
| AD Meds/ CCW Non-AD Dementia (VA EMR) | 1.09 (1.08,1.10) | 16.12 | 1.77E-58 |
| AD Meds/CCW Non-AD -Dementia (VA EMR) 60+ | 1.10 (1.09,1.11) | 17.41 | 7.04E-68 |

| | | | |
|---|---|---|---|
| AD Meds/ CCW Non-AD Dementia (VA EMR) 65+ | 1.10 (1.09,1.11) | 17.63 | 1.25E-69 |
| AD Meds/ MVP-Dementia (VA EMR) | 1.07 (1.06,1.09) | 15.03 | 4.40E-51 |
| AD Meds/ MVP-Dementia (VA EMR) 60+ | 1.09 (1.08,1.10) | 16.43 | 1.11E-60 |
| AD Meds/ MVP-Dementia (VA EMR) 65+ | 1.09 (1.08,1.10) | 16.78 | 3.33E-63 |
| AD Meds/ MVP-ADRD (VA EMR) | 1.10 (1.08,1.11) | 16.87 | 8.05E-64 |
| AD Meds/ MVP-ADRD (VA EMR) 60+ | 1.11 (1.09,1.12) | 18.01 | 1.51E-72 |
| AD Meds/ MVP-ADRD (VA EMR) 65+ | 1.11 (1.10,1.13) | 18.37 | 2.22E-75 |
| AD Meds/ Phe-Dementia (VA EMR) | 1.09 (1.08,1.11) | 16.97 | 1.37E-64 |
| AD Meds/ Phe-Dementia (VA EMR) 60+ | 1.11 (1.09,1.12) | 18.10 | 3.16E-73 |
| AD Meds/ Phe-Dementia (VA EMR) 65+ | 1.11 (1.10,1.12) | 18.35 | 3.38E-75 |
| AD Meds/ MVP- AD+ (VA EMR) | 1.10 (1.09,1.11) | 17.01 | 6.79E-65 |
| AD Meds/ MVP- AD+ (VA EMR) 60+ | 1.11 (1.10,1.12) | 18.30 | 7.71E-75 |
| AD Meds/ MVP- AD+ (VA EMR) 65+ | 1.12 (1.11,1.13) | 18.65 | 1.33E-77 |
| AD Meds/ CCW-AD (VA EMR) | 1.11 (1.10,1.13) | 16.64 | 3.62E-62 |
| AD Meds/ CCW-AD (VA EMR) 60+ | 1.13 (1.11,1.14) | 18.04 | 9.69E-73 |
| AD Meds/ CCW-AD (VA EMR) 65+ | 1.13 (1.12,1.15) | 18.26 | 1.68E-74 |
| AD Meds/ Phe-AD (VA EMR) | 1.11 (1.10,1.13) | 16.53 | 2.27E-61 |
| AD Meds/ Phe-AD (VA EMR) 60+ | 1.13 (1.11,1.14) | 17.84 | 3.62E-71 |
| AD Meds/ Phe-AD (VA EMR) 65+ | 1.13 (1.12,1.15) | 18.05 | 7.30E-73 |
| AD Meds/ MVP-AD (VA EMR) | 1.11 (1.10,1.13) | 16.66 | 2.69E-62 |
| AD Meds/ MVP-AD (VA EMR) 60+ | 1.13 (1.11,1.14) | 17.96 | 3.65E-72 |
| AD Meds/ MVP-AD (VA EMR) 65+ | 1.13 (1.12,1.15) | 18.08 | 4.61E-73 |

**Supplemental Table 6.** Performance of each algorithm compared to the chart review outcomes of "Likely Not" versus "Possible/Likely" AD (A) and Dementia (B) with VA EMR data and VA EMR/CMS data.

| | Algorithm | Sensitivity | Specificity | Positive Predictive Value (PPV) | Negative Predictive Value (NPV) |
|---|---|---|---|---|---|
| **A) Compared to Chart Review AD** | MVP-AD (VA EMR) | 0.71 (0.60,0.81) | 0.95 (0.90,0.96) | 0.90 (0.79,0.96) | 0.85 (0.78,0.90) |
| | MVP-AD (VA EMR/CMS) | 0.80 (0.70,0.89) | 0.87 (0.79,0.92) | 0.78 (0.67,0.87) | 0.88 (0.81,0.93) |
| | CCW-AD (VA EMR) | 0.80 (0.70,0.89) | 0.92 (0.86,0.96) | 0.86 (0.76,0.93) | 0.89 (0.82,0.93) |
| | CCW-AD (VA EMR/CMS) | 0.88 (0.79,0.94) | 0.84 (0.77,0.90) | 0.77 (0.67,0.85) | 0.92 (0.86,0.96) |
| | Phe-AD (VA EMR) | 0.71 (0.60,0.81) | 0.95 (0.90,0.96) | 0.90 (0.79,0.96) | 0.85 (0.78,0.90) |
| | Phe-AD (VA EMR/CMS) | 0.82 (0.71,0.90) | 0.87 (0.79,0.92) | 0.78 (0.68,0.87) | 0.89 (0.82,0.94) |
| | MVP-AD+ (VA EMR) | 0.96 (0.89,0.99) | 0.62 (0.53,0.71) | 0.60 (0.51,0.69) | 0.96 (0.90,0.99) |
| | MVP-AD+ (VA EMR/CMS) | 0.97 (0.91,1.00) | 0.55 (0.46,0.64) | 0.56 (0.48,0.65) | 0.97 (0.90,1.00) |
| | MVP-ADRD (VA EMR) | 0.97 (0.91,1.00) | 0.57 (0.48,0.65) | 0.57 (0.48,0.66) | 0.97 (0.91,1.00) |
| | MVP-ADRD (VA EMR/CMS) | 0.97 (0.91,1.00) | 0.50 (0.41,0.59) | 0.54 (0.45,0.63) | 0.97 (0.89,1.00) |
| | AD-Med (VA EMR) | 0.75 (0.65,0.84) | 0.70 (0.61,0.78) | 0.60 (0.49,0.70) | 0.82 (0.74,0.89) |
| | AD-Med (VA EMR/CMS) | 0.75 (0.65,0.84) | 0.70 (0.61,0.78) | 0.60 (0.49,0.70) | 0.82 (0.74,0.89) |
| | AD-Med/Phe-AD (VA EMR) | 0.87 (0.82,0.96) | 0.69 (0.60,0.76) | 0.62 (0.52,0.71) | 0.90 (0.82,0.95) |
| | AD-Med/Phe-AD (VA EMR /CMS) | 0.91 (0.82,0.96) | 0.66 (0.57,0.74) | 0.62 (0.52,0.71) | 0.92 (0.85,0.97) |
| | AD-Med/MVP-AD (VA EMR) | 0.87 (0.77,0.94) | 0.69 (0.60,0.76) | 0.62 (0.52,0.71) | 0.90 (0.82,0.95) |
| | AD-Med/MVP-AD (VA EMR/CMS) | 0.91 (0.82,0.96) | 0.66 (0.57,0.74) | 0.62 (0.52,0.71) | 0.92 (0.85,0.97) |
| | AD-Med/CCW-AD (VA EMR) | 0.89 (0.80,0.95) | 0.69 (0.60,0.76) | 0.63 (0.53,0.72) | 0.92 (0.84,0.96) |
| | AD-Med/CCW-AD (VA EMR/CMS) | 0.92 (0.84,0.97) | 0.66 (0.57,0.74) | 0.62 (0.52,0.71) | 0.93 (0.86,0.98) |
| | AD-Med/MVP-AD+ (VA EMR) | 0.99 (0.93,1.00) | 0.50 (0.41,0.59) | 0.54 (0.45,0.62) | 0.98 (0.92,1.00) |
| | AD-Med/MVP-AD+ (VA EMR/CMS) | 0.99 (0.93,1.00) | 0.46 (0.37,0.55) | 0.52 (0.45,0.62) | 0.98 (0.91,1.00) |
| | AD-Med/MVP-ADRD (VA EMR) | 0.99 (0.93,1.00) | 0.46 (0.38,0.56) | 0.52 (0.44,0.61) | 0.98 (0.91,1.00) |
| | AD-Med/MVP-ADRD (VA EMR/CMS) | 0.99 (0.93,1.00) | 0.42 (0.33,0.51) | 0.50 (0.42,0.59) | 0.98 (0.90,1.00) |
| **B) Compared to Chart Review Dementia** | MVP-ADRD (VA EMR) | 0.92 (0.86,0.96) | 0.81 (0.71,0.89) | 0.88 (0.82,0.93) | 0.86 (0.77,0.93) |
| | MVP-ADRD (VA EMR/CMS) | 0.94 (0.89,0.98) | 0.75 (0.64,0.84) | 0.85 (0.78,0.91) | 0.89 (0.79,0.96) |
| | MVP-Dementia (VA EMR) | 0.94 (0.88,0.97) | 0.66 (0.54,0.76) | 0.81 (0.75,0.94) | 0.87 (0.75,0.94) |

| | | | | |
|---|---|---|---|---|
| MVP-Dementia (VA EMR/CMS) | 0.95 (0.90,0.98) | 0.58 (0.47,0.69) | 0.78 (0.71,0.84) | 0.88 (0.77,0.96) |
| CCW-Non-AD Dementia (VA EMR) | 0.74 (0.66,0.82) | 0.81 (0.71,0.89) | 0.86 (0.78,0.92) | 0.67 (0.56,0.76) |
| CCW-Non-AD Dementia (VA EMR/CMS) | 0.80 (0.72,0.87) | 0.73 (0.62,0.83) | 0.82 (0.75,0.89) | 0.70 (0.59,0.79) |
| Phe-Dementia (VA EMR) | 0.90 (0.83, 0.94) | 0.77 (0.66,0.86) | 0.86 (0.79,0.92) | 0.82 (0.72,0.90) |
| Phe-Dementia (VA EMR/CMS) | 0.94 (0.89,0.98) | 0.67 (0.56,0.77) | 0.82 (0.75,0.88) | 0.88 (0.77,0.95) |
| AD-Med (VA EMR) | 0.65 (0.55,0.73) | 0.81 (0.71,0.89) | 0.84 (0.75,0.91) | 0.59 (0.49,0.69) |
| AD-Med (VA EMR/CMS) | 0.65 (0.55,0.73) | 0.81 (0.71,0.89) | 0.84 (0.75,0.91) | 0.59 (0.49,0.69) |
| AD-Med/MVP-ADRD (VA EMR) | 0.95 (0.90,0.98) | 0.68 (0.57,0.78) | 0.83 (0.75,0.88) | 0.90 (0.79,0.96) |
| AD-Med/MVP-ADRD (VA EMR/CMS) | 0.97 (0.92,0.99) | 0.63 (0.52,0.74) | 0.81 (0.73,0.87) | 0.93 (0.82,0.98) |
| AD-Med/MVP-Dementia (VA EMR) | 0.97 (0.92,0.99) | 0.54 (0.43,0.66) | 0.77 (0.70,0.83) | 0.91 (0.80,0.98) |
| AD-Med/MVP-Dementia (VA EMR/CMS) | 0.98 (0.93,0.99) | 0.48 (0.37,0.60) | 0.75 (0.67,0.81) | 0.93 (0.80,0.98) |
| AD-Med/CCW-Non-AD-Dementia (VA EMR) | 0.86 (0.79,0.92) | 0.68 (0.57,0.78) | 0.81 (0.73,0.87) | 0.76 (0.64,0.85) |
| AD-Med/ CCW-Non-AD-Dementia (VA EMR/CMS) | 0.90 (0.83,0.94) | 0.63 (0.52,0.74) | 0.79 (0.72,0.86) | 0.79 (0.67,0.89) |
| AD-Med/Phe-Dementia (VA EMR) | 0.94 (0.88,0.97) | 0.66 (0.54,0.76) | 0.81 (0.74,0.87) | 0.87 (0.75,0.94) |
| AD-Med/Phe-Dementia (VA EMR/CMS) | 0.97 (0.92,0.99) | 0.57 (0.45,0.68) | 0.78 (0.71,0.84) | 0.92 (0.80,0.98) |

**Reviewer Guide**

### MVP MCI/AD/Dementia Project Chart Review Guide

- The goal of this chart review project is to create a “gold standard” to use as a benchmark for comparing different automated diagnoses of age-related cognitive disorders from electronic medical record (EMR) data based on ICD code algorithms, machine learning, and other techniques.
- We are working to create standardized classifications of subjects as having mild cognitive impairment (MCI), Alzheimer’s disease (AD), and dementia.
- Given the nature of the VA medical record, the type of evidence you would need to decide with 100% certainty that a patient had an age-related cognitive disorder (i.e., MCI, AD, or dementia) will likely not be available. As such, we are proposing that subjects be classified as “Likely”, “Possible”, or “Likely not” for having had each of the conditions. For example, *Likely AD*, *Possible AD*, or *Likely not AD*. These classifications are italicized throughout this guide to emphasize that these are categories that we have defined and to contrast them to the actual conditions when applicable.
- Note that none of the guidelines presented here are prescriptive, and are here only to orient the chart reviewer to the type of evidence available in the medical record and the parameters of our study. There will be situations where the available evidence will not fit nicely into one of the described categories above, or which are on the border between levels of certainty. Please use your best judgement in these cases.

*In the chart review, you will be asked to (1) identify indicators of three conditions (MCI, AD, and Dementia) and (2) classify subjects as “Likely”, “Possible”, or “Likely not” for having had each of the three conditions. A description of the three conditions follows:*

**Mild Cognitive Impairment (MCI):** As mild cognitive impairment is often a stage encountered on the way to a dementia diagnosis, an MCI case would be a subject who has indications of a period of reduced cognitive function. It would *not* include people whose first sign of cognitive difficulty in the medical record is severe enough to warrant a full dementia diagnosis, although presumably there may have been a period of time, if brief, between normal function and dementia. We also note that this is conceptualized as a degenerative condition which often leads to dementia. Therefore, the trajectory of cognitive functioning can be informative. That is, subjects whose evidence for impairment is low but who subsequently develop a dementia can be classified as *Possible MCI* or *Likely MCI* cases, even in the absence of cognitive screening data such as MMSE scores. Conversely, someone who has a period of poor cognition, even with documented low cognitive screening scores, but who subsequently has a return to normal cognition could be classified as *Likely not MCI*. However, that said, the subsequent development of dementia should not be considered a necessary condition of being a *Likely MCI* case. As cognitive ability can vary in the population, a decline in functioning from previous levels may be more indicative of cognitive impairment than absolute performance on a test.

**Alzheimer's Disease (AD):** AD is a progressive form of dementia with impairment in memory and judgement. It is the most common form of dementia. Definitive diagnosis of AD is made at autopsy based on neuropathological criteria, but this sort of definitive indicator will not be available for most patients in the VA EMR. Therefore, the determination of *Likely AD* has to be made based on the examinations available in the medical record. Strong evidence such as a full diagnostic work-up by a neurologist or in a memory clinic and/or an MRI with signs consistent with AD pathology are enough for a *Likely AD* diagnosis unless subsequent notes indicate additional complicating factors. Note that signs of other pathology such as vascular problems are not necessarily disqualifying, but indications that cognitive impairment is due to some other significant problem such as medication issues could be. Note that VA clinicians are often reluctant to use an AD diagnosis or ICD code, and instead assign a non-specific dementia code. Repeated use of these non-specific terms or codes should not be considered evidence against an AD diagnosis.

**Dementia**: Dementia is a broad term used to refer to any of a cadre of disorders and diseases which can impair function in domains such as memory, judgement, and language. In addition to AD, other forms of dementia include Vascular dementia, Lewy Body dementia, Huntington's Disease, Parkinson's Disease Dementia, Frontotemporal/Other Fronto Dementia, Normal Pressure Hydrocephalus, and Korsakoff Syndrome. It also includes rare disorders such as Pick's disease and Creutzfeldt-Jakob Disease. In many cases, insufficient evidence may be available in the medical record to confirm that someone specifically has AD, but the level of impairment is sufficient to confirm that a subject definitely had some form of dementia. In that case, a person might be a *Possible AD* case, but a *Likely Dementia* case. In general, as AD is a form of dementia, the reported level of confidence for a dementia diagnosis must be at least as high as their level of AD diagnosis certainty, such that someone who is classified as *Possible AD*, must be classified as either *Possible Dementia* or *Likely Dementia*.

Indications for the different levels of confidence – *Likely*, *Possible*, and *Likely not*:

**MCI:**

*Likely MCI*

- The strongest evidence for MCI is likely to be evidence of a positive screen for memory problems and/or poor cognitive testing by a specialist, with scores consistent with mild impairment. A *Likely MCI* rating is also achievable after examining the total of evidence from multiple indicators in the absence of other transitory mitigating factors.
- Indicators of MCI:
    - Documentation of an impaired score on a screening test (MMSE, MoCA, or SLUMS) given in a general clinic.
    - Evidence of AD medication for someone without a full diagnosis for dementia.
    - Report of a decline in functioning from previous levels by patient self-report or report of an adult son or daughter, partner, or close friend.
    - Mention of an MCI diagnosis given outside of the VA healthcare system.
    - Subsequent progression to dementia after a period of reported or substantiated mild impairment.
    - Some clinicians use the term "Mild AD" to describe MCI; judgement may be required in these instances to determine whether the condition is truly MCI versus an AD/dementia process.

*Possible MCI*

- One or more indicators above can be enough to indicate a *Possible MCI* diagnosis, as long as these are not attributable to some other factor (e.g. medication).

*Likely not MCI*

- A subject with none of the indicators of MCI above.
- A subject with cognitive difficulties traced to another cause.
- A dementia case without any documentation of a period of reduced cognitive function prior to dementia onset should be classified as *Likely not MCI.*
- As subjects can have a "bad day," subjects with a transitory period of reduced cognitive function, even including a low test on a cognitive screening score (MMSE, MoCA, SLUMS), who subsequently remits and possibly tests in the normal range and remains there for the remaining portion of the medical record, should be classified as *Likely not MCI.*

**AD:**

*Likely AD*

- The clearest indicator in the medical record of *Likely AD* is screening for AD symptomatology in a specialty clinic such as neurology, gerontology, or neuropsychology, or a memory-specific clinic. (At some VAs, this screening is often carried out in Psychiatry/Geropsychiatry or Mental Health clinics). Often the report states the diagnosis or findings as "consistent with AD;" this may be the strongest endorsement of AD given prior to autopsy. When available, an MRI evaluation consistent with AD and/or post-mortem confirmation would also be strong evidence. In the absence of screening in a specialty clinic, a classification of *Likely AD* is appropriate when the preponderance of evidence points to the subject as having AD, including multiple indicators of AD from the list below.
- Indicators of AD:
    - Reports of AD diagnosis outside of the VA healthcare system.
    - Reports of AD screening and treatment in general practice clinics, including performance on cognitive screeners consistent with AD conducted outside of the formal testing completed in specialty clinics as described above.
    - Use of AD medication. Note, however, that often medications for AD are offered to subjects with subjective memory complaints and without strong evidence of AD-specific pathology, so, in general, report of AD medications should not be considered strong evidence of the presence of AD. However, based on the cost, Memantine (Namenda) often requires an additional layer of review before prescription in the VA, so treatment with Memantine should be weighted as greater evidence for the presence of AD than other AD medications.
    - Participation in an AD treatment study.
    - Symptom progression consistent with the gradual cognitive decline characteristic of dementia, especially if paired with eventual placement in an inpatient residential facility.
    - MRI scan reporting pathology consistent with AD, even in the presence of vascular pathology.
        - Note: Selecting "abnormal" means *consistent with AD*. This may require a judgment call if not specified in the MRI report.
        - If there is an "abnormal" finding on CT/MRI but it does not inform or contribute to your diagnostic impression/decision for AD—for example, generalized atrophy or a bullet wound—then select "normal".
            - It may be helpful to ask yourself, "Does the imaging support the diagnosis below?" If it does, select "abnormal".

*Possible AD*

- *Possible AD* may be given when no specialist evaluation has been performed, but when several indicators of AD are present, but not at the level that would be sufficient to conclude *Likely*

*AD*.

- The *Possible AD* category might also be used when there is a potentially confounded explanation for dementia symptomatology. Note that in some cases, uncertainty in the type of dementia makes it appropriate for a patient to be classified as *Possible AD* but also *Likely Dementia* (see below for guidance on dementia diagnosis).
    - Example: If AD is specifically listed as part of a differential diagnosis, *Possible AD* is warranted. **However, if a patient is diagnosed with "Mixed Dementia" and AD is one of the specified diagnoses, this should be coded as *Likely AD*.**

*Likely not AD*

- This is the category for subjects without indicators of AD or for which evidence is so minimal that a *Possible AD* case is not appropriate.

Other Things to Note:

- Often, diagnoses are copy/pasted into the EMR by one clinician based on prior entries, such that multiple listings of the same diagnosis should not be taken as additional evidence of the diagnosis unless new screening, evaluation, or treatment for AD/dementia is obtained in the visit.
- In general, co-occurring conditions are not exclusionary for AD; for example, stroke may occur in someone with AD. However, when a co-occurring condition represents a possible alternate explanation, this should be considered carefully and a *Possible AD* classification may be more appropriate.
- The results of a comprehensive neuropsychological evaluation should trump a mention of AD (or multiple mentions of AD) in the notes. For example, if there are several mentions of AD throughout the notes, but a subsequent neuropsychological assessment concludes that the pattern of results is *not* consistent with AD, this would be coded as *Likely not AD*.

**Dementia**:

*Likely Dementia*

- Similar to *Likely AD*, the clearest indicator in the medical record of *Likely Dementia* is a full evaluation at a specialty clinic such as neurology, gerontology, or neuropsychology, or a memory-specific clinic. (At some VAs, this screening is often carried out in Psychiatry/Geropsychiatry or Mental Health clinics). When available, an MRI evaluation consistent with dementia and/or post-mortem confirmation would also be strong evidence. *Likely Dementia* would include any *Likely AD* cases and cases of any other specific dementias. *Likely Dementia* would also include subjects who have a clear dementia process, but for whom the specific type of dementia is not clear. Many of the indicators of dementia mirror those for AD.
- Indicators of Dementia:
    - Reports of dementia diagnosis outside of the VA healthcare system.

- Reports of dementia screening and treatment in general practice clinics, including performance on cognitive screeners consistent with dementia conducted outside of the formal testing completed in specialty clinics as described above.
- Use of AD medication. Note, however, that often medications for AD are offered to subjects with subjective memory complaints and without strong evidence of AD-specific pathology.
- Participation in a dementia treatment study.
- Symptom progression consistent with dementia.
- MRI scan reporting pathology consistent with dementia.
  - Note: Selecting "abnormal" means *consistent with dementia*. This may require a judgment call if not specified in MRI report.
  - If there is an "abnormal" finding on CT/MRI but it does not inform or contribute to your diagnostic impression/decision for dementia—for example, generalized atrophy or a bullet wound—then select "normal".
    - It may be helpful to ask yourself, "Does the imaging support the diagnosis below?" If it does, select "abnormal".

*Possible Dementia*

- This category would be for subjects with one or more of the above indicators, but without enough evidence to meet a *Likely Dementia* threshold. This may be due to abnormal presentation, lack of documentation, or the presence of potential confounding factors.

*Likely not Dementia*

- Like the *Likely not* categories for MCI and AD, most subjects in this category will be patients without any documentation of dementia in the medical record. This category may also include subjects who have an indicator of dementia, but insufficient documentation to conclude *Possible Dementia* or disease course inconsistent with dementia (e.g., short-term remitted period of delirium).

*Missing Value*

There is one other possible rating which could be assigned to each patient, which is a *Missing Value*. As conceived here, the *Likely not MCI/AD/Dementia* categories include subjects without any documentation of one of these disorders. You should be able to make a classification into one of the three levels (*Likely, Possible, Likely not)* for the majority of subjects. The *Missing Value* category should be used sparingly, and only in cases where evidence is so contradictory to make any sort of definitive statement difficult to impossible. This could be, for example, when evidence of cognitive impairment has only occurred in conjunction with other disorders (e.g. schizophrenia) or in cases where test results are so contradictory such that it is not clear that valid measurements of cognitive performance and state have been obtained. In most cases, uncertainty can be incorporated into the *Possible* level of confidence.

Thank you for your participation!
